# Surgery & COVID-19: A rapid scoping review of the impact of COVID-19 on surgical services during public health emergencies

**DOI:** 10.1101/2020.12.03.20243592

**Authors:** Connor M O’Rielly, Joshua S Ng-Kamstra, Ania Kania-Richmond, Joseph C Dort, Jonathan White, Jill Robert, Mary Brindle, Khara M Sauro

## Abstract

**Background:** Healthcare systems globally have been challenged by the COVID-19 pandemic, necessitating the reorganization of surgical services to free capacity within healthcare systems.

**Objectives:** To understand how surgical services have been reorganized during and following public health emergencies, and the consequences of these changes for patients, healthcare providers and healthcare systems.

**Methods:** This rapid scoping review searched academic databases and grey literature sources to identify studies examining surgical service delivery during public health emergencies including COVID-19, and the impact on patients, providers and healthcare systems. Recommendations and guidelines were excluded. Screening was completed in partial (title, abstract) or complete (full text) duplicate following pilot reviews of 50 articles to ensure reliable application of eligibility criteria.

**Results:** One hundred and thirty-two studies were included in this review; 111 described reorganization of surgical services, 55 described the consequences of reorganizing surgical services and six reported actions taken to rebuild surgical capacity in public health emergencies. Reorganizations of surgical services were grouped under six domains: case selection/triage, PPE regulations and practice, workforce composition and deployment, outpatient and inpatient patient care, resident and fellow education, and the hospital or clinical environment. Service reorganizations led to large reductions in non-urgent surgical volumes, increases in surgical wait times, and impacted medical training (i.e., reduced case involvement) and patient outcomes (e.g., increases in pain). Strategies for rebuilding surgical capacity were scarce, but focused on the availability of staff, PPE, and patient readiness for surgery as key factors to consider before resuming services.

**Conclusions:** Reorganization of surgical services in response to public health emergencies appears to be context-dependent and has far-reaching consequences that must be better understood in order to optimize future health system responses to public health emergencies.

**ARTICLE SUMMARY:** *Strengths and limitations of the study:*

- This rapid scoping review provides an exhaustive and rigorous summary of the academic and grey literature regarding modifications to surgical services in response to public health emergencies, especially COVID-19.
- This study did not limit studies based on location or language of publication to ensure a worldwide pandemic had contributions from worldwide voices.
- Both quantitative and qualitative outcomes were included, with a mix of inductive and deductive data abstraction approaches to provide a comprehensive understanding of surgical services during public health emergencies.
- Studies with potential relevance to this question are emerging at an unprecedented rate in response to the COVID-19 pandemic and as such, some may not be included in the current review.

*Original protocol for the study:* As requested, the original unpublished protocol for this study is included as a supplementary file.

*Funding statement:* This study did not receive grant from any funding agency in the public, commercial or not-for-profit sectors.

*Competing interest statement:* All authors declare that they have no competing interests in accordance with the International Committee of Medical Journal Editors uniform declaration of competing interests.

## INTRODUCTION

The novel SARS-CoV-2 (COVID-19) virus has spread across the globe with unrelenting speed. At the time of writing, over 16 million cases have been confirmed with 650,000 fatalities.^1^ To protect the most vulnerable in our societies, efforts to curb further escalation (e.g., travel restrictions, physical distancing) have had a focal objective: to prevent surges that could overwhelm healthcare including shortages in personal protective equipment (PPE), ventilators, and hospital capacity.

Medical institutions have taken steps to maximize staff, PPE, ventilators, and intensive care unit (ICU) capacity in case public health efforts to ‘flatten the curve’ are insufficient. Most notably, surgical programs have suspended non-urgent (or ‘elective’) surgical procedures. Non-urgent surgeries are often defined as procedures for which a delay of three months or longer would not result in significant adverse effects to the patient.^2 3^ These changes have thrust patients, providers, and healthcare organizations into previously unexplored territory.

While governing bodies such as colleges and academies of surgery have made recommendations to alter surgical service delivery in response to COVID-19, they have not always provided explicit instructions on how programs should operationalize the recommendations. As such, approaches to surgical triage and service delivery remain unclear: who has done what where, and why? Further, the impacts of adopting these recommendations on surgical programs, and more importantly, the physical and psychological well-being of patients and healthcare providers have only been hypothesized.^4^ Lastly, as COVID-19 begins to release its grip on the world and the post-pandemic recovery begins, programs will be tasked with rebuilding the surgical capacity necessary to reschedule and resume the backlog of postponed procedures. Evidence distilled from the experiences of others in the context of COVID-19 and other public health emergencies (i.e., H1N1, Ebola, SARS) is needed to guide approaches to surgical service delivery.

To enable evidence-informed reorganization and resumption of non-urgent surgeries post-COVID-19 and for future public health emergencies, we conducted a rapid scoping review to synthesize relevant and available literature. Our aim was to understand how surgical services were reorganized in response to COVID-19 and other public health emergencies; how reorganization impacted patients, healthcare providers, and health systems; and what approaches have been taken to resume surgical service delivery.

## METHODS

### Study Design

This scoping review followed the Joanna Briggs Institute methodology and Preferred Reporting Items for Systematic reviews and Meta-Analysis extension for Scoping Reviews (PRISMA-ScR) checklist.^5 6^ The rapidly evolving situation of the current COVID-19 pandemic demanded a similarly rapid evidence synthesis. Therefore, methodological concessions recommended by the World Health Organization and Cochrane guidance for rapid reviews were made.^7 8^ This review addressed three questions: 1) How have surgical services been reorganized in response to public health emergencies? 2) What are the patient-, healthcare provider-, and system-level consequences of reorganizing surgical services? and 3) What approaches were used for resuming surgical services?

### Search Strategy

The search strategy was developed by two investigators (CO, KS) and refined by others with context expertise in surgery and literature review methodology (JNK, AKR). The search strategy included subject headings, keywords, and synonyms identifying public health emergencies in general and specific public health emergencies (Ebola, SARS-CoV1, H1N1, MERS), and surgery; and were tailored for each database (Appendix A). Given the exploratory nature of the review we did not filter by study design or publication type, and since the impacts of a pandemic spans many countries there were no language restrictions.

We used the search strategy to search MEDLINE (including Epub Ahead of Print, In-Process & Other Non-Indexed Citations) and Embase from inception until May 8, 2020. Anticipating pertinent information may not be published (i.e., joint statements, recommendations, and guidelines from surgical colleges) we supplemented the database search with a structured grey literature search including targeted website searching, advanced and general Google searching, and contact with knowledge experts (Appendix A).^9^ The reference lists of included studies were screened for relevant studies not otherwise captured.

### Study Selection

Titles and abstracts were reviewed by one of two independent reviewers with a third, independent reviewer screening 25% of randomly selected references in duplicate. Full texts of studies considered potentially eligible at title/abstract screening phase by at least one reviewer were reviewed in duplicate by two reviewers for eligibility. Any disagreement between reviewers at the full text screening phase was resolved through discussion and did not necessitate a third reviewer. If studies were excluded at the full text screening phase, the reason for exclusion was noted. Full text articles meeting eligibility criteria were included and data were abstracted using a standardized data abstraction form (Appendix B). At both stages of screening, a pilot sample of 50 articles were jointly reviewed by both reviewers to ensure reliable application of eligibility criteria between reviewers.

### Study Eligibility

Studies were eligible for inclusion if they discussed alterations to surgical services during public health emergencies and reported: 1) reorganization of surgical services, 2) impact of reorganizing surgical services on patients, healthcare providers, or healthcare system or 3) approaches to resuming surgical capacity. Studies of any design or publication date were eligible for inclusion. Studies in any language were eligible, but consistent with rapid review methods, studies not easily translated by authors were excluded from the data synthesis, although citations are still provided. Studies were excluded if they described: only urgent interventions arising during a hospital admission (e.g., emergency tracheostomy, caesarean section), settings beyond in-patient acute care (e.g., outpatient clinics including dental clinics), surgical services beyond a public health emergency without comparison to public health emergencies, healthcare services not specifically related to surgical service.

Notably, our intention was to include guidelines that made recommendations regarding provision of surgical services; however, a high-quality review of guidelines was published^10^ during the preparation of this review and as such, we chose to exclude guidelines.

### Data Extraction

Data were abstracted by one reviewer, and verified by a second reviewer, using a standardized data abstraction form (Appendix B). Data included: date of publication, country, study design, definition of non-urgent surgery, characteristics of study sample (if applicable), outcomes of interest for the three research questions, detailed below.

### Outcomes of Interest

Our primary outcomes were reorganization of surgical services, impact of reorganization and resuming surgical services. We intentionally included a broad array of outcomes and used an inductive approach to data abstraction to gain a comprehensive understanding of surgical services and the impact during public health emergencies.

We collected qualitative data from studies reporting on changes to surgical programming, conceptualized into five categories: changes to triage criteria or case selection, changes to PPE practices, workforce changes, changes to patient care, changes to resident and fellow education, and environmental changes. Qualitative and quantitative data on the impact of reorganization of surgical services was organized by impact on: patients, providers and healthcare system. To illustrate temporal changes, data preceding, during and after the precipitating event were collected whenever possible. Quantitative variables of interest included: adverse events (including morbidity and mortality), primary care and emergency department visits, number of hospital and ICU admissions, length of hospital and ICU stay, number of surgical procedures performed and number of procedures cancelled, care costs, and wait times for non-urgent surgery. Qualitative variables included narrative description of patient or physician experience, written descriptions of changes to physician remuneration, or comments surrounding surgical waitlist composition. Qualitative data was also collected on details of efforts to rebuild capacity to surgical services.

### Study Quality (Risk of Bias) Assessment

Given the aim of a rapid scoping review is not to appraise evidence but to map the available literature,^11^ quality appraisal of included studies was not performed.

### Data Synthesis, Analysis and Reporting

Study and sample data were summarized narratively in a data table and using descriptive statistics where appropriate. We decided *a priori* to use a random effects model for meta-analysis if there was sufficient data on the impact of changes to surgical services to pool, however this was not feasible. Instead, descriptive statistics were used to synthesize the quantitative outcomes. Data were synthesized and presented separately for each of the three research questions.

### Patient and Public Partnership

Patients and the public were not involved in the conception or analysis of this review.

## RESULTS

### Search Results

A total of 3 013 unique scholarly articles and 106 sources of grey literature were identified, of which 702 were considered eligible for full text review. After full text review, 120 studies and five documents from the grey literature were included. Screening of the reference lists of included articles led to seven additional studies being included for a total of 132 included studies. Thirty-seven studies contributed data to more than one of the research questions resulting in the qualitative synthesis of 111 studies assessing alterations to service delivery, 55 studies evaluating the consequences of these changes, and six studies enumerating their procedures for rebuilding capacity (Table 1). The flow of evidence sources within the study is detailed in Figure 1. One Spanish language study was translated for inclusion,^12^ but two studies could not be readily translated therefore they are not included in the synthesis.^13 14^

**Table 1.**
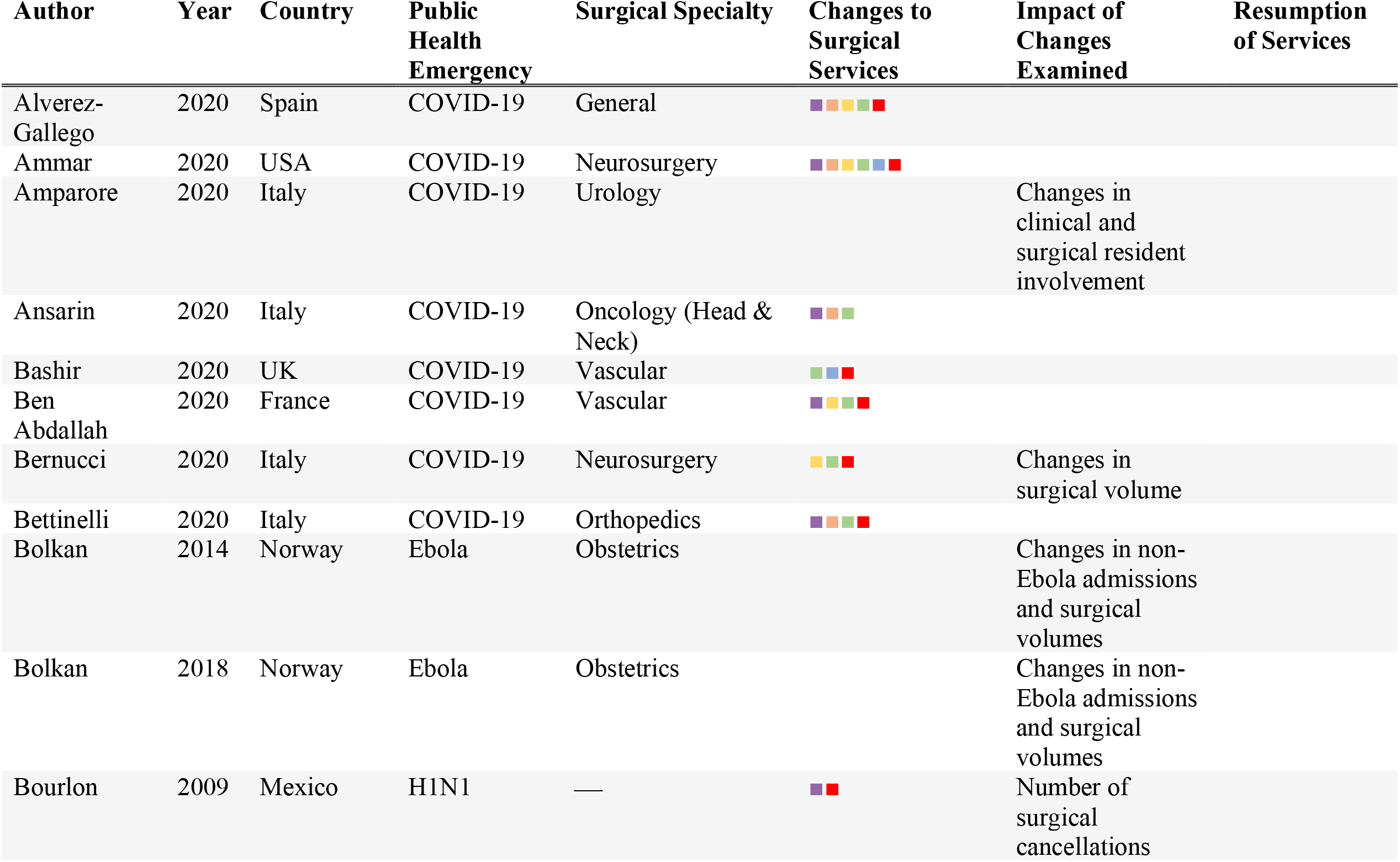
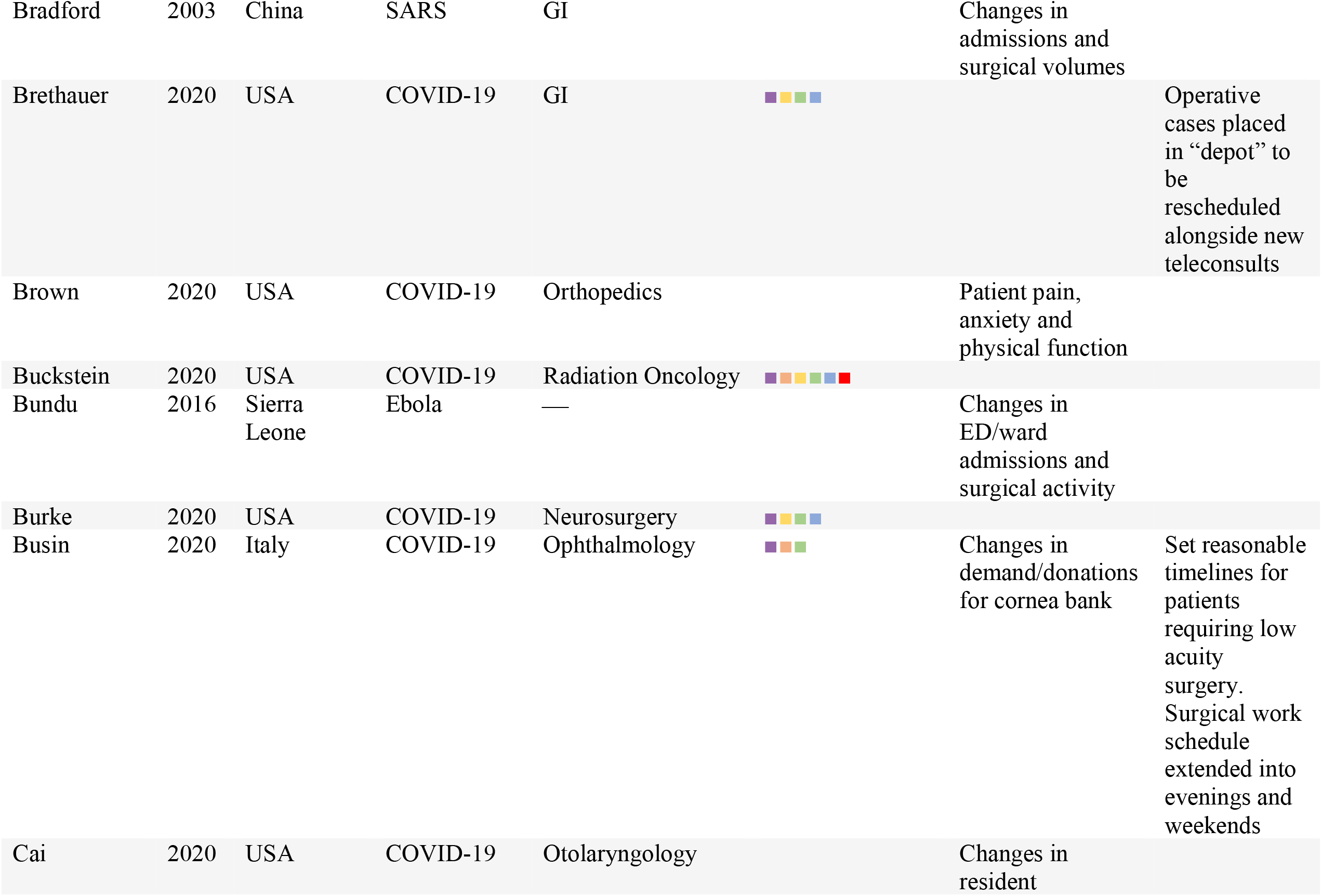
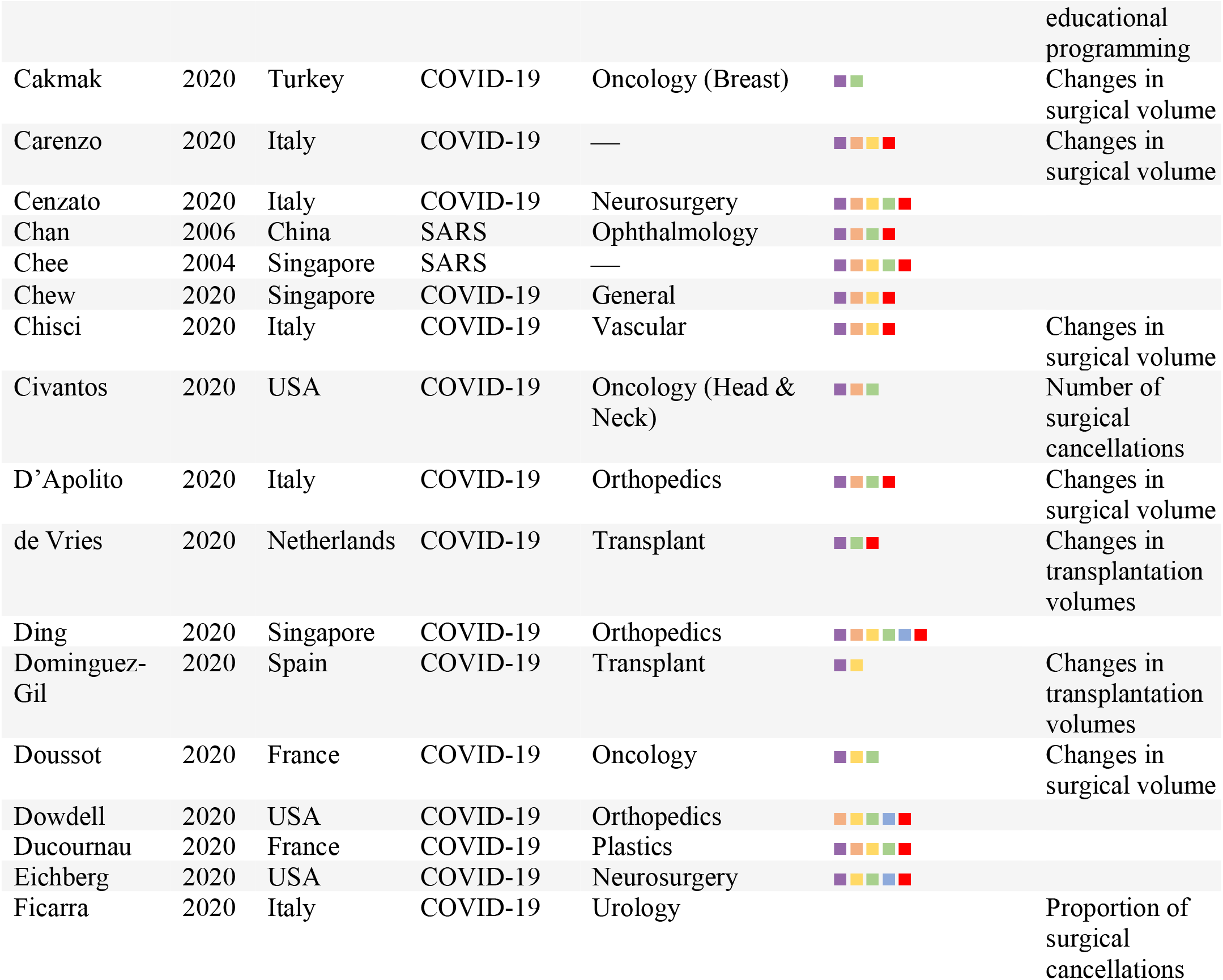
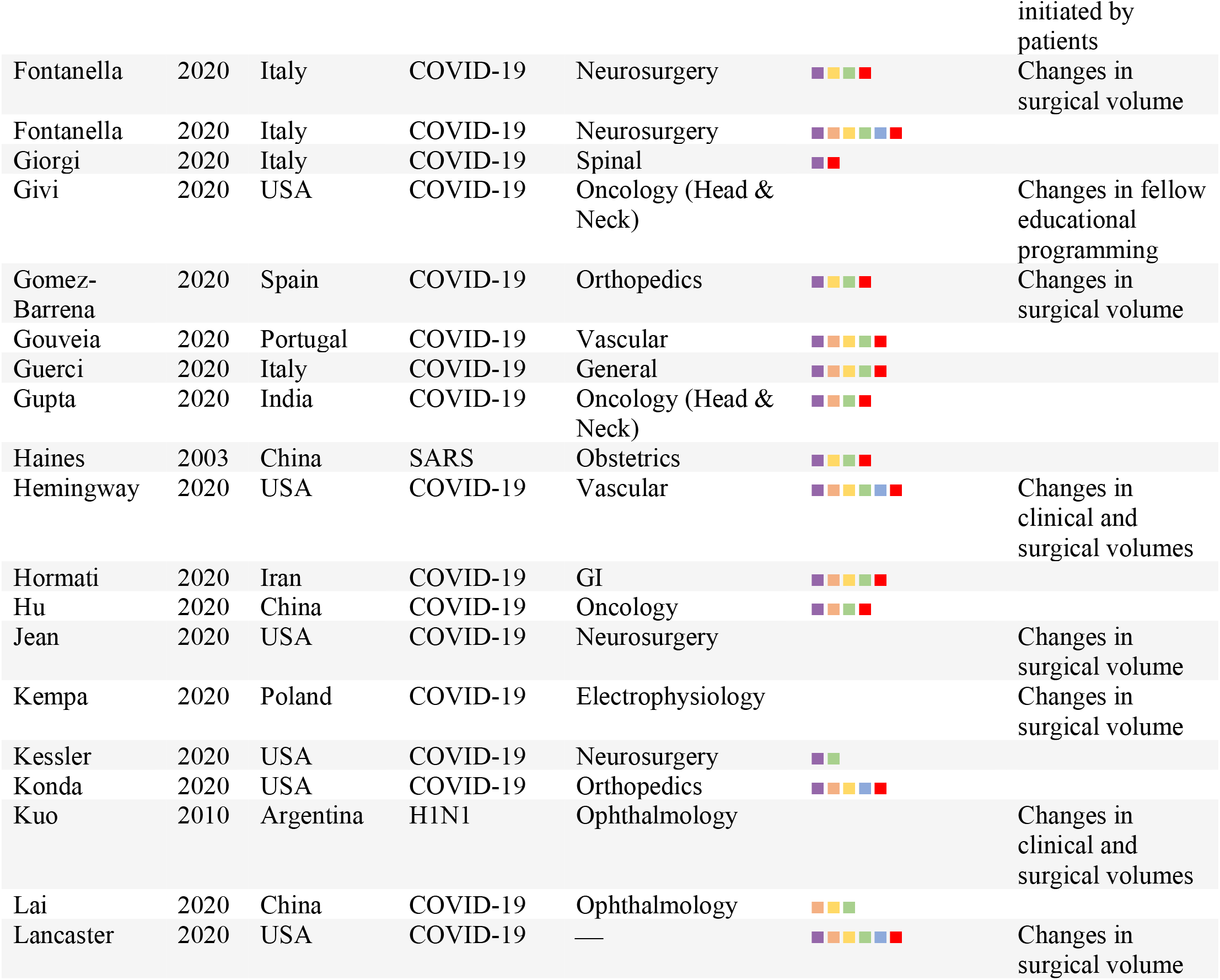
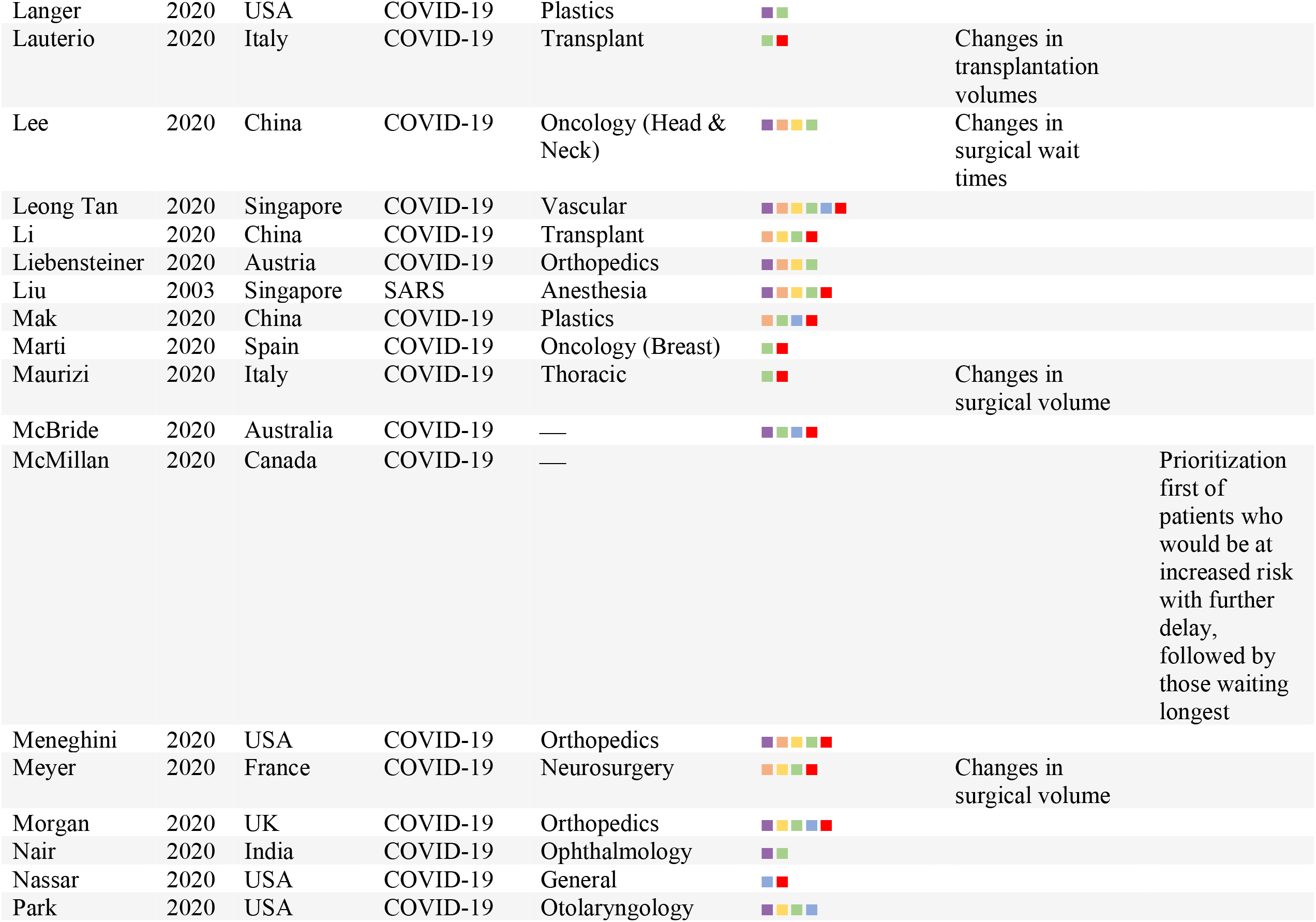
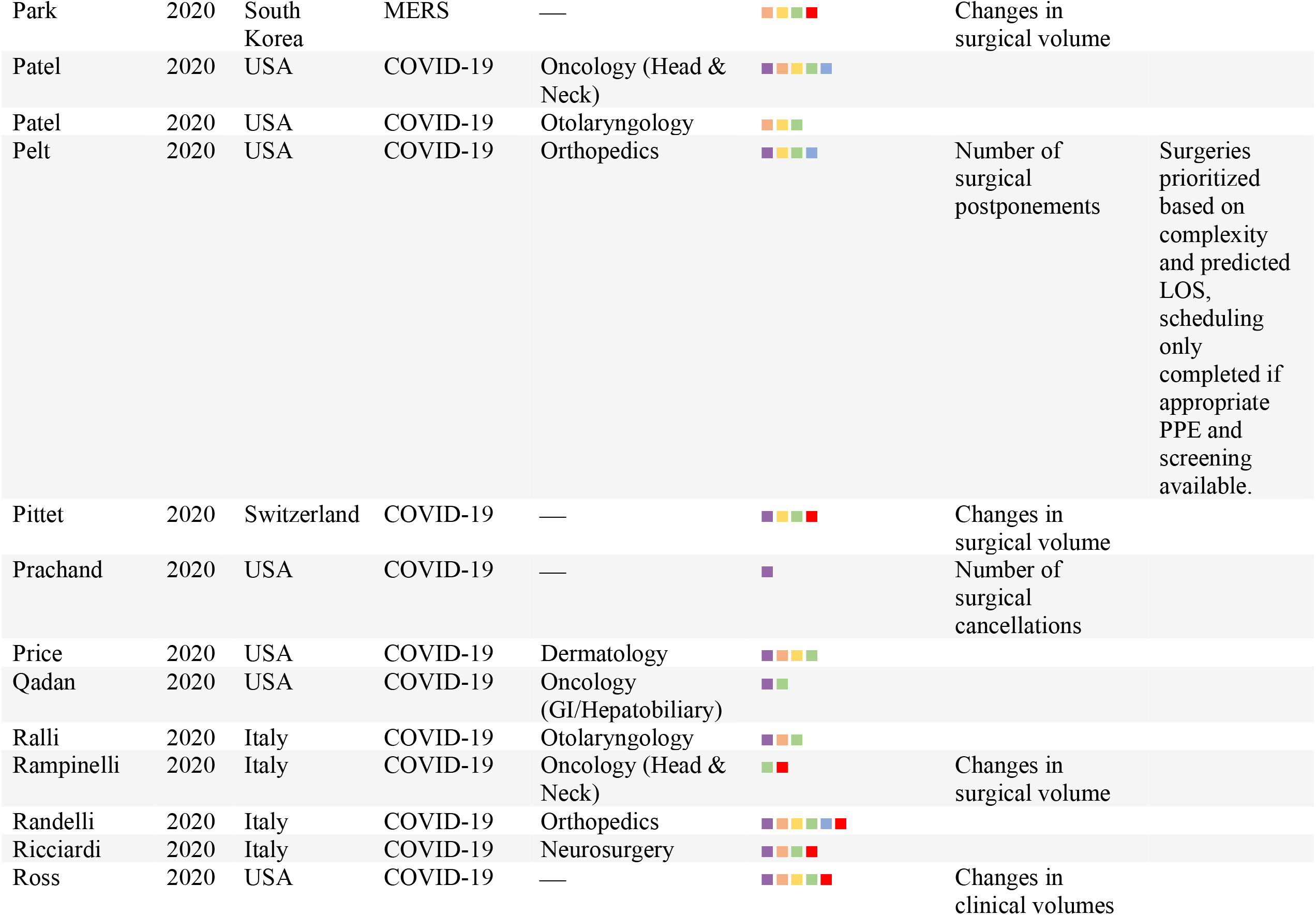
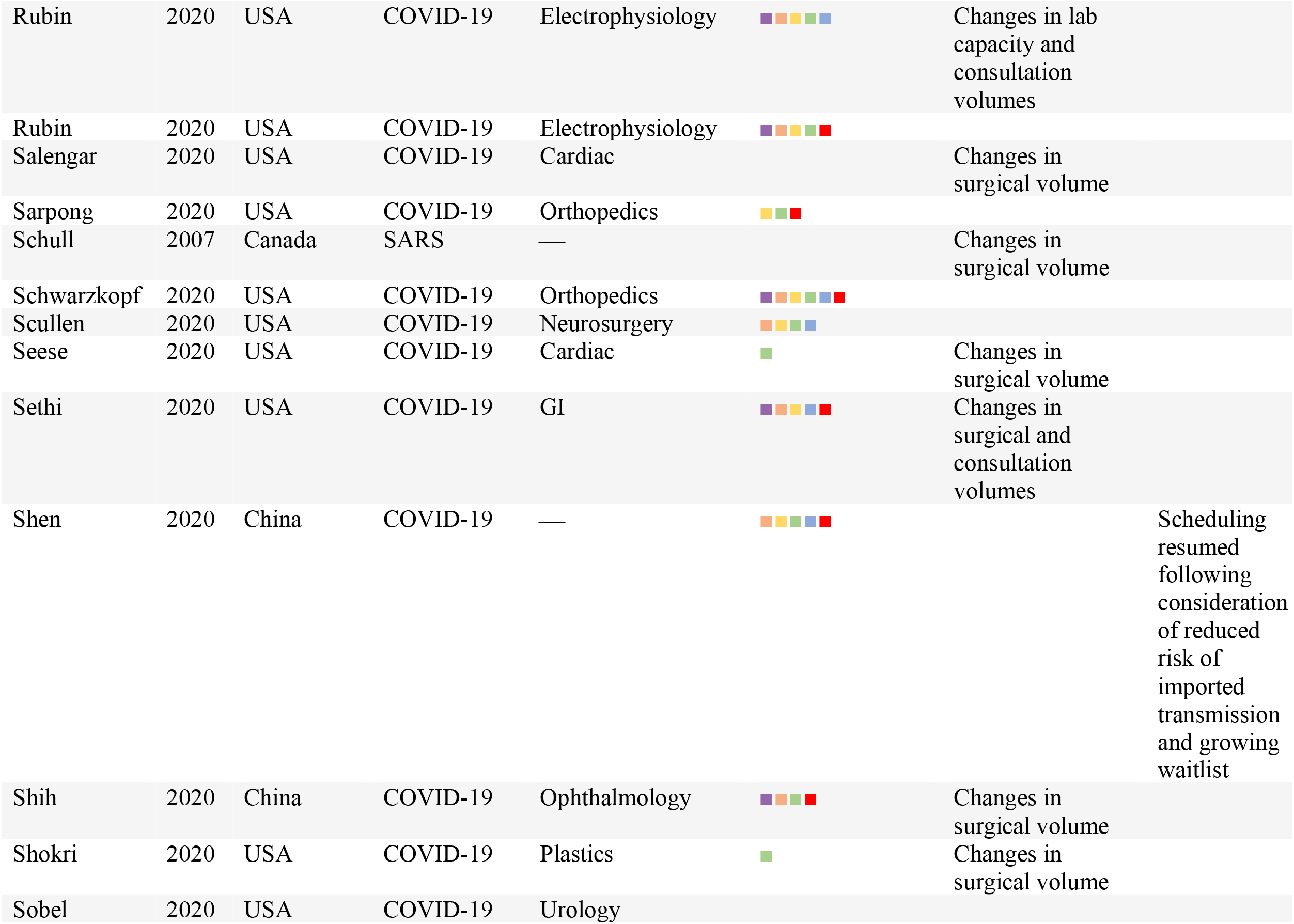
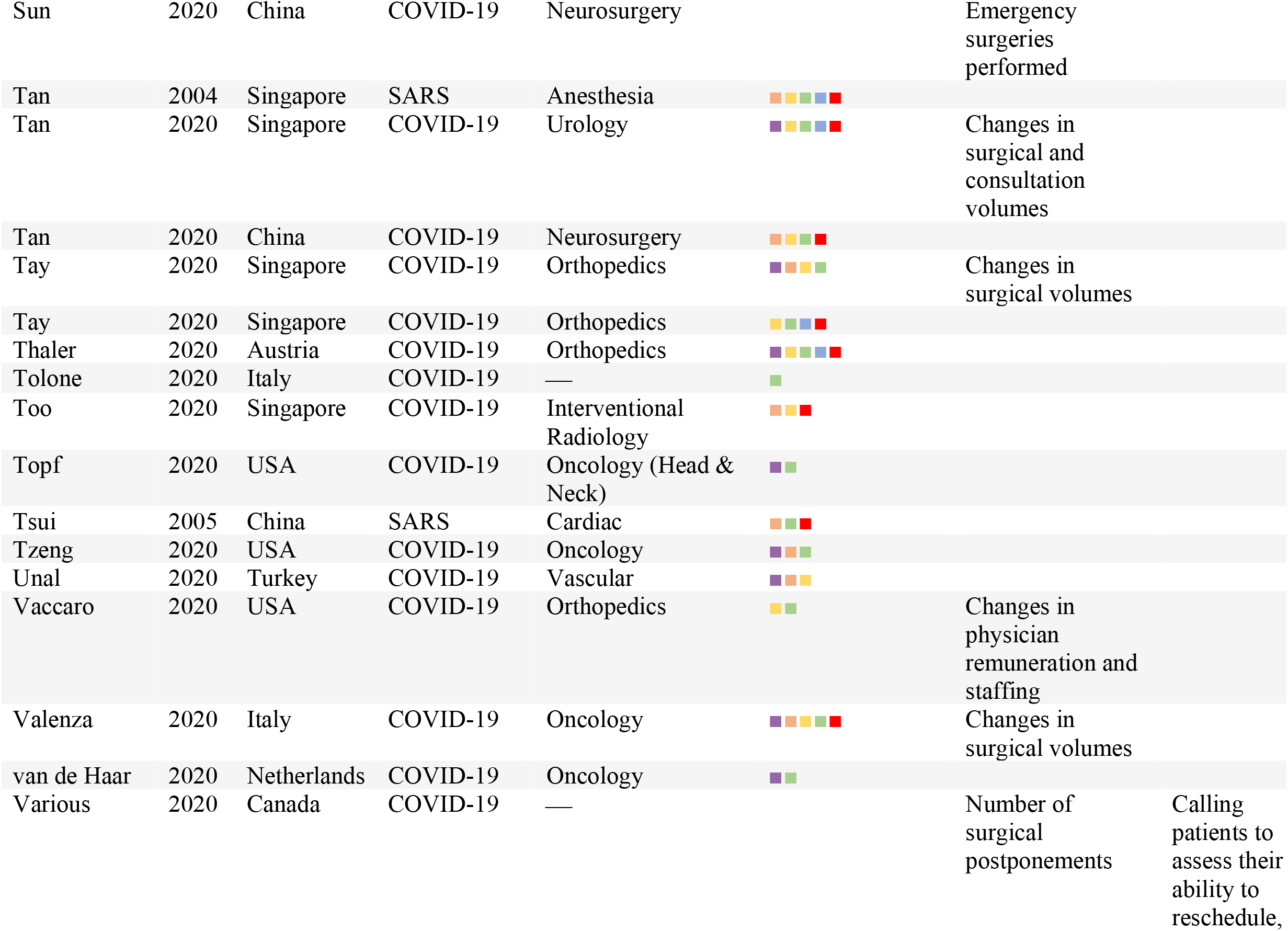
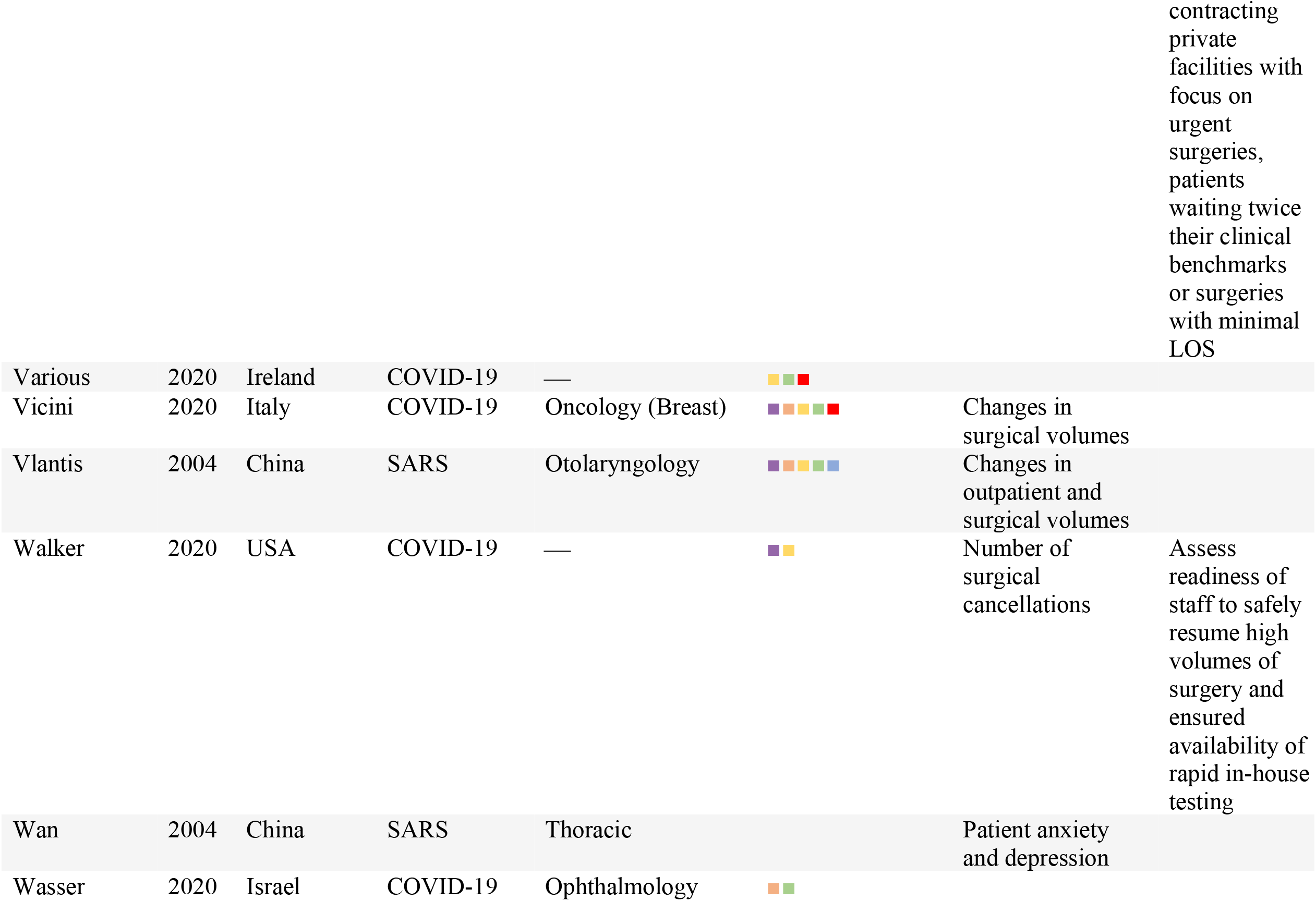
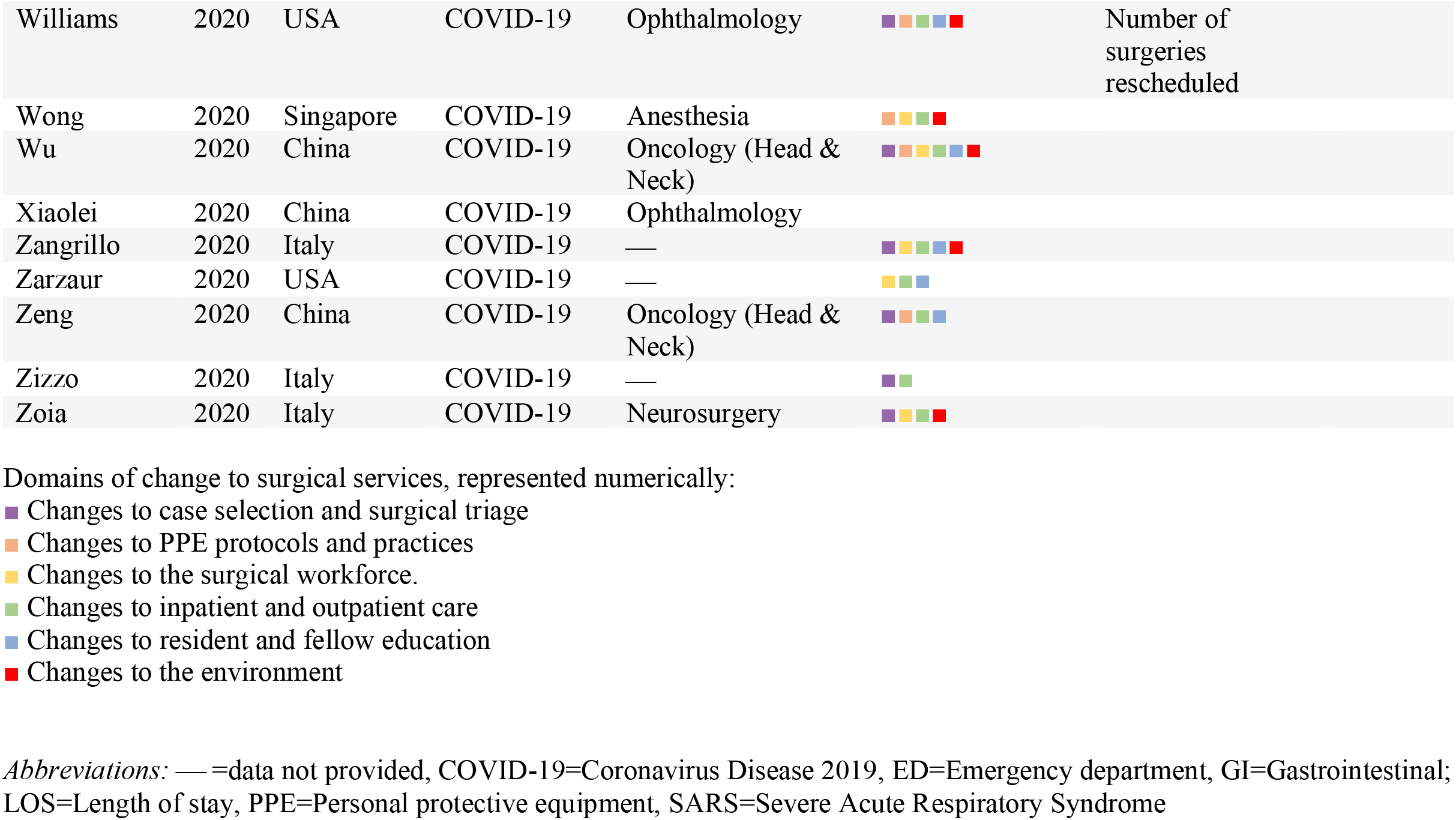
Description of included studies

**Figure 1.**
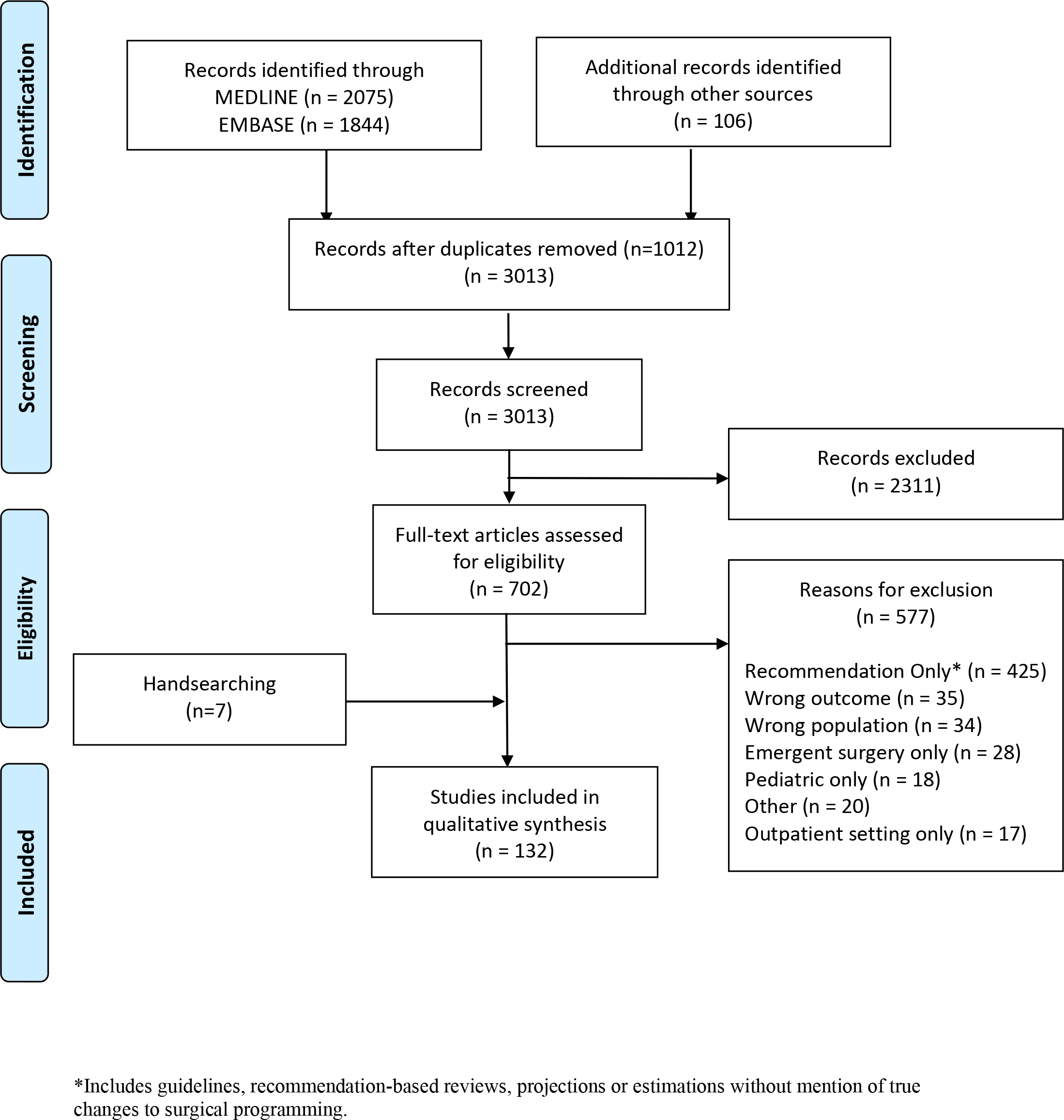
Flow of studies in the scoping review. *Includes guidelines, recommendation-based reviews, projections or estimations without mention of true changes to surgical programming.

### Description of Studies

The majority of included studies were published in 2020 about COVID-19 (87.9%, n=116); fewer studies were related to other public health emergencies: SARS (7.58%, n=10), Ebola (2.27%, n=3), H1N1(1.52%, n=2), and MERS (0.76%, n=1). Over two thirds of the included studies (74.2%) emerged from the countries hit earliest by COVID-19; China (14.4%, n=19), Singapore (8.33%, n=11), Italy (19.7%, n=26), and the USA (31.8%, n=41). While many studies described the experiences of their surgical departments as a whole, oncology (15.9%, n=21), orthopedics (13.6%, n=18), and neurosurgery (11.4%, n=15) were the specialties most prominently represented. Summaries of descriptive study information are shown in Figure 2.

**Figure 2.**
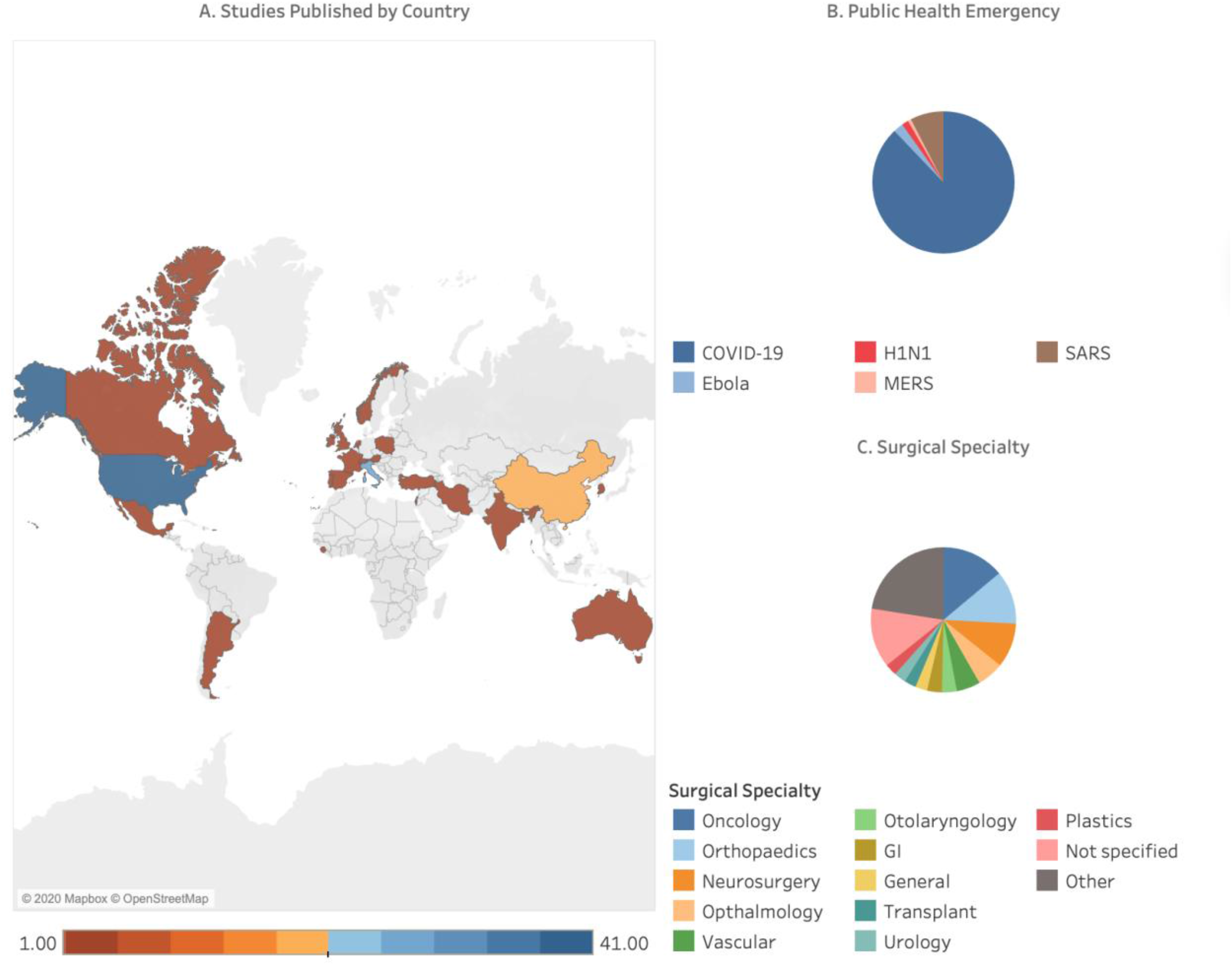
Study characteristics. **A**. Country of publication, **B**. Public health emergency discussed, and **C**. Surgical specialty addressed (‘Other’ includes Cardiac (n=3), Anesthesia (n=3), Electrophysiology (n=3), Obstetrics and Gynecology (n=3), Thoracic (n=2), Interventional Radiology (n=1), and Dermatology (n=1)).

### Reorganization of Surgical Service

A number of themes emerged from the 108 studies describing reorganization of surgical services. Nearly all studies reported partial, with most reporting full cessation of non-urgent surgeries at their centre, albeit with varying definitions of “non-urgent” (e.g., can be safely postponed for 3 months) and “urgent” (e.g., patient would have adverse outcome if not completed within 7 days). Changes to service delivery were focused on six domains: case selection/triage, PPE regulations and practice, workforce composition and deployment, outpatient and inpatient patient care, resident and fellow education, and the hospital or clinical environment (Table 2). The three domains that were most frequently reported (case selection/triage, patient care, and workforce) are described in greater detail below.

**Table 2.**
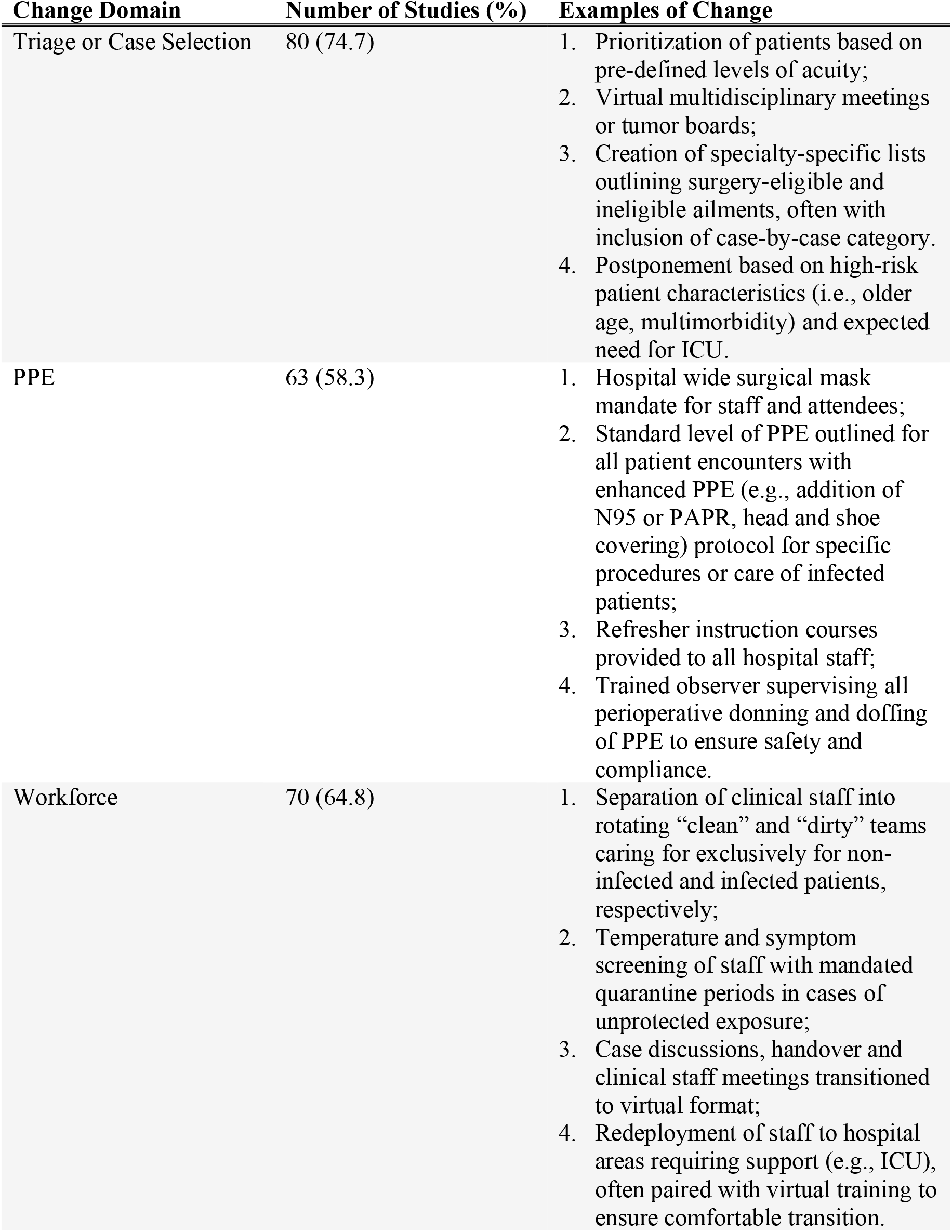
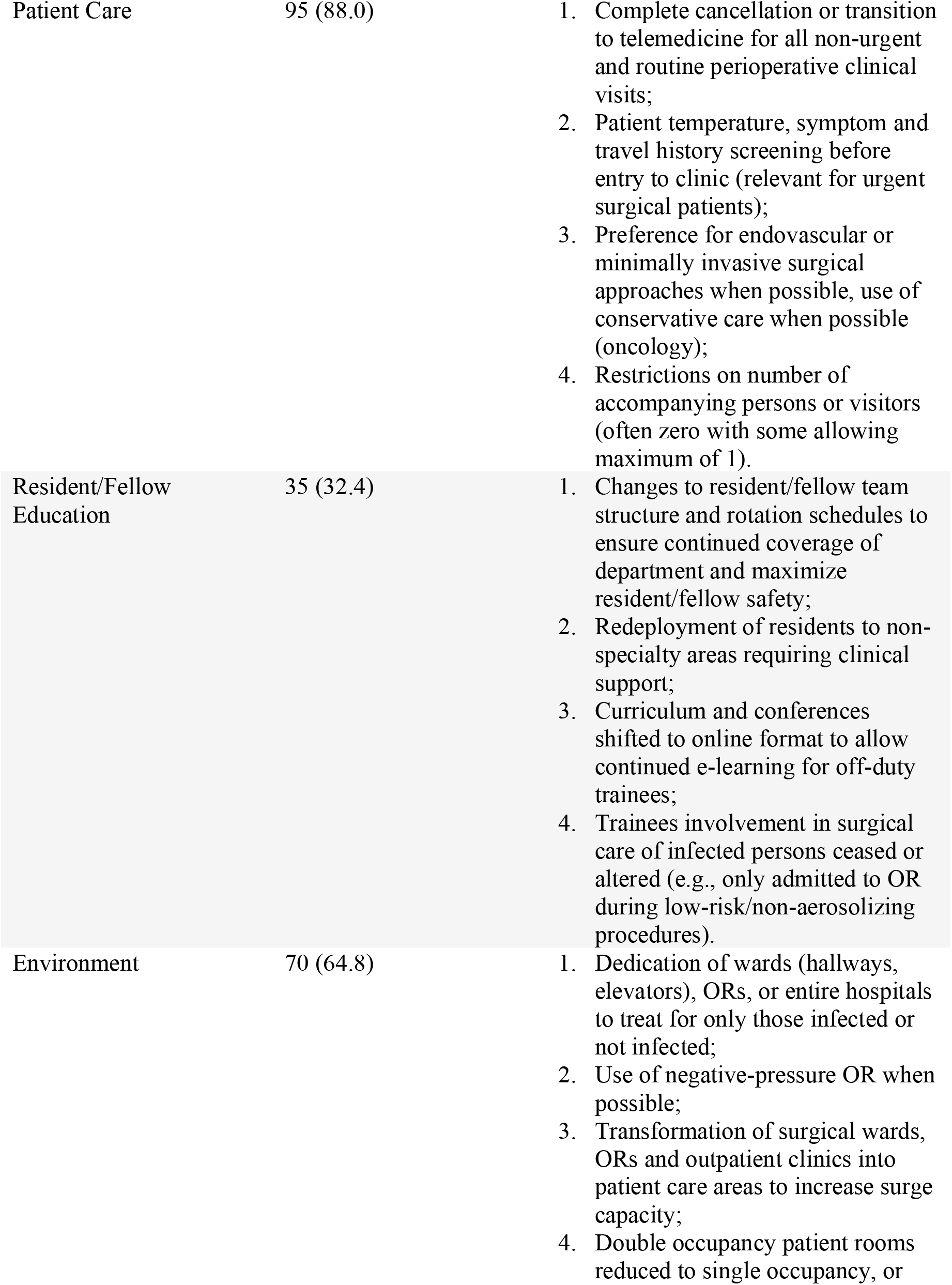
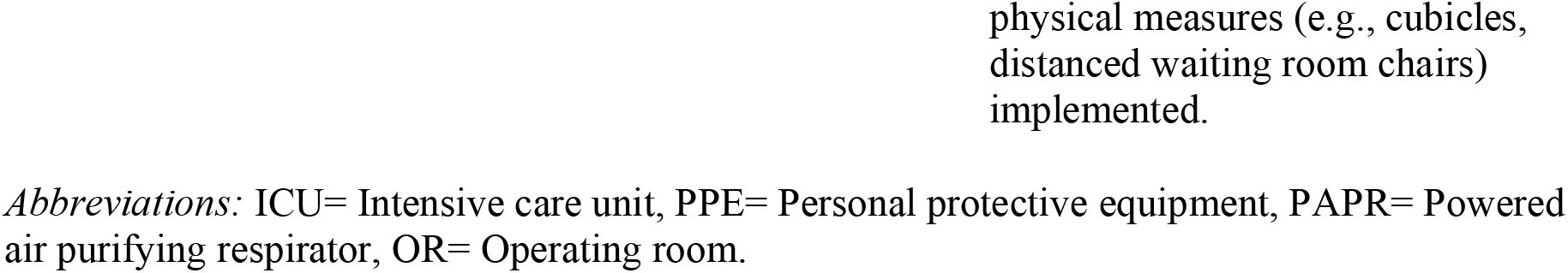
Reorganization of surgical services, by domain.

#### 1. Changes to Case Selection and Triage Procedures

The “where” of surgery is described above; however, the issue of what patients can safely undergo which surgical procedures was also discussed in the included studies. We identified cancelling or postponing “non-urgent” surgeries was almost universal. Most often hospitals cancelled surgeries via telephone or text message, but some studies identified that patients initiated their own surgical cancellations due to concerns with safety and contamination. While urgent surgeries were triaged according to routine practice, new triage decisions were made for non-urgent (including oncology) procedures. Methods for triaging non-urgent procedures varied across studies, from the use of guideline supported checklists of eligible procedures to virtual multidisciplinary meetings where the treating surgeon presented details of the case (e.g., patient characteristics, acuity, imaging) to a larger group representing many surgical specialties to reach consensus on each case.

#### 2. Changes to Patient Care

Sixty-two studies reported complete cessation or marked reduction of in-person, non-urgent outpatient clinic visits. In these studies, only urgent patients and those requiring post-operative suture or staple removal were granted in-person visits under strict conditions including mask wearing, negative symptom check, history or temperature pre-screening. Studies specific to COVID-19 almost universally filled the resulting care gap for patients deemed “non-urgent” using telephone or video-based telemedicine. Interfaces used include, but were not limited to Zoom, WeChat, Facetime, telephone, and SMS text messaging. A reported advantage of telemedicine was the ability to not only follow-up with returning patients but to also continue consultations and establish contact with new patients who would require care when non-urgent surgeries resumed. While some admitted a historical reluctance to transition to video-based telemedicine and reported early concerns with their ability to establish secure connections with patients, frequently their worries faded with use and many reported telemedicine would remain integrated in their practices beyond the pandemic.

#### 3. Changes to the Workforce

Fourteen of the included studies describe changing the workforce into a minimum of two teams; a “contaminated” team providing care to infected patients and a “clean” team managing those not infected. When these teams were kept separate from one another both inside and outside of the hospital setting, surgical departments were able to continue managing the inevitable emergencies (as well as non-urgent procedures in some settings) without cross contamination during the public health emergencies. New work rotations and shift schedules were created to ensure this structure was sustainable, often with extra healthcare providers designated to replace those with exposures and to provide adequate time off to prevent burnout. This practice was only possible with wards, operating rooms, and pathways (i.e. corridors, elevators) that are separated under the same “clean” and “contaminated” designation. In the most extreme case, entire hospitals were designated for each patient group, as was done by Singapore during SARS^15^ and Italy during COVID-19.^16^

### Impact of Reorganizing Surgical Services

Of the 55 studies with data relevant to this question, 42 were focused on changes in surgical volumes with six reporting changes to surgical waitlist time or composition, four underlining changes to resident and fellow involvement in surgery, and two showing changes in patient pain, anxiety, and depression. These recurring outcome measures are summarized below with data for all studies relevant to this question shown in Appendix C.

#### Changes in Surgical Volumes

Thirty-seven studies provided data for this outcome, with 37.8% (n=14) reporting a greater than 75% reduction and 70.3% (n=26) reporting a greater than 50% reductions in their overall or site specific non-urgent surgical volumes (Figure 3a). Not all studies reported reductions; as one study from an oncology “hub” hospital in Italy reported a 20% increase in their surgical volumes, likely due to more cases being diverted to their hospital during the COVID-19 pandemic.^17^

**Figure 3.**
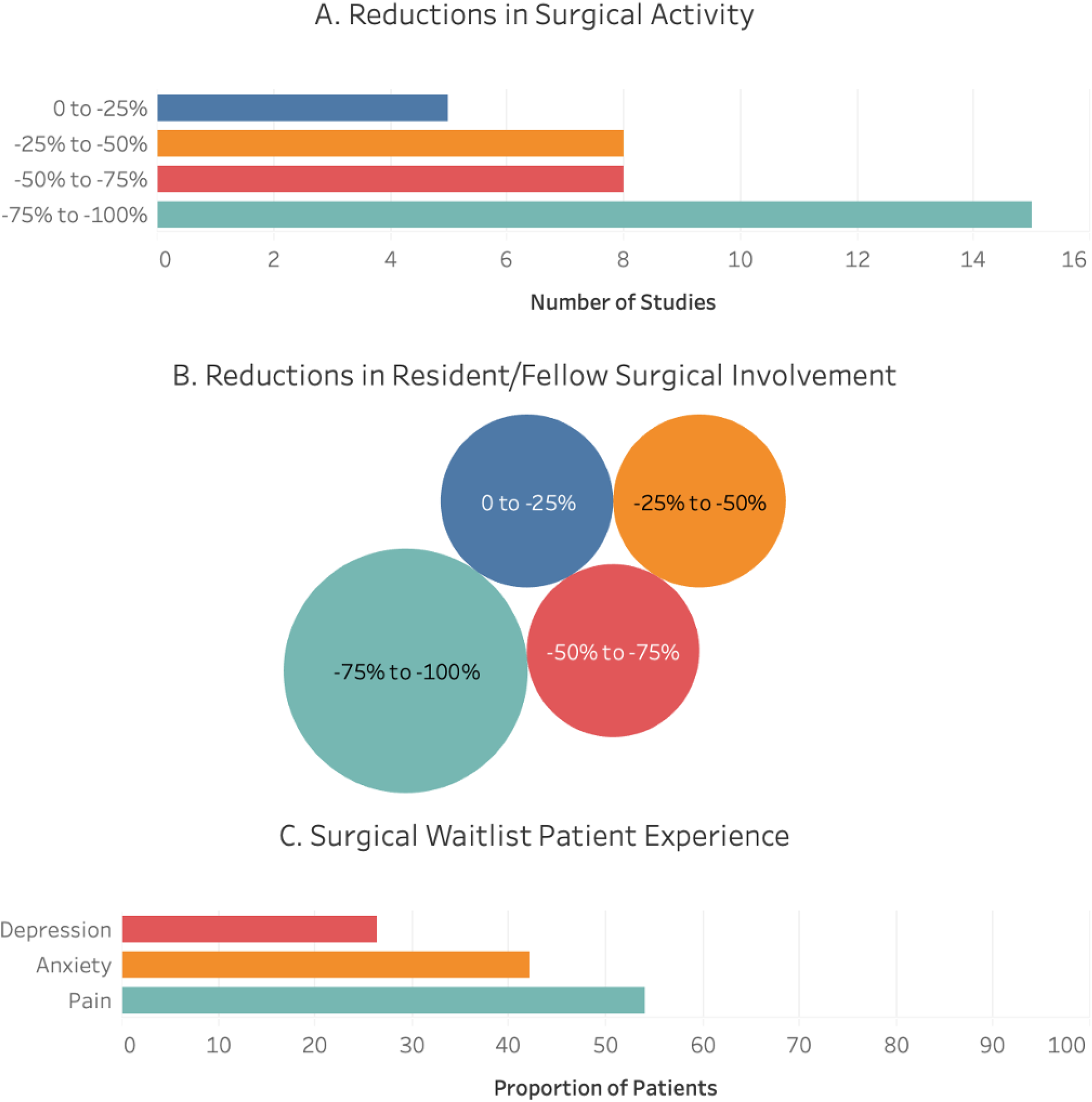
Summary of the impacts of alterations to surgical services. A summary of the impacts of alterations to surgical services during public health emergencies on **A**. Overall surgical activity (n=37 studies), **B**. Resident and fellow involvement in surgery (n=5 studies) where circle size represents the number of studies contributing to that quartile, and **C**. Patient experience (n=2 studies).

#### Changes in Resident/Fellow Involvement in Surgical Activities

Four studies^18-21^ reported on this outcome; two survey-based case series, one resident-level case study and one study containing both survey and case log data. The reductions in surgical involvement for residents are shown by quartile in Figure 3b.

#### Changes to Waitlist Length and Composition

Five studies ^22-26^ reported data for this outcome. One centre reported a 64% increase in length of their minor colorectal surgery waitlist^25^ and another centre (head and neck oncological surgery program) reported a 500% increase in latency from diagnosis to surgery.^26^ One study reported no waitlist deaths during the COVID-19 pandemic^24^ while another saw a small decrease in the number of weekly waitlist deaths.^23^ A single study identified more patients leaving their renal transplantation waitlist due to mortality or clinical deterioration.^22^

#### Changes in Patient Pain, Anxiety, and Depression

Two studies^27 28^ reported pain, anxiety, and depression among more than half of waitlist patients; 42.1% experienced anxiety, and 26.3% experienced depression (Figure 3c). The leading reported cause of patient anxiety was a lack of knowledge about when their surgeries would be rescheduled. Other than a single study describing the negative financial effects of the COVID-19 pandemic,^29^ impacts on healthcare providers and their practices were rarely discussed.

### Rebuild Surgical Capacity

A total of seven studies reported the experience of rebuilding surgical capacity in their departments, hospitals, or systems; all studies referred to the COVID-19 pandemic. One study from China reported reopening non-urgent surgeries with close consideration of risk for imported transmission but did not provide further detail of triage or prioritization.^30^ Among studies that changed their surgical triage practices, patients were prioritized for surgery based on procedure acuity or urgency (i.e., risk to patients if surgery were further delayed), resource intensity, and procedural complexity. Four studies^31-34^ noted that prior to resuming non-urgent surgeries, availability of the staff, (Operating Room) ORs, PPE, and testing was necessary to prepare for a large and complicated surgical backlog.

## DISCUSSION

This review identified over 3,000 evidence sources, 132 of which were included. Approaches to reorganizing surgical services varied between studies and centers, but the cancellation or postponement of non-urgent surgeries such as arthroplasty surgeries for chronic joint pain, coronary artery bypass graft surgery for asymptomatic individuals, and primary gastric bypass surgery was nearly universal.^2^ The most frequently reported change to surgical services was modified triage criteria for surgical cases, workforce, and approach to patient care. Many studies reported a decrease in surgical volumes due to public health emergencies, while a few reported the non-surgical impacts such as patient wellbeing or changes in healthcare utilization beyond the surgical wards. Very few studies described their experience resuming surgical services after a public health emergency.

The varied approaches to providing surgical services during a public health emergency identified in this review illustrate that a “one fits all” approach does not exist. Changes to surgical services likely depends on the characteristics of specific centers and their patients. While several guidelines have been published with recommendations on how to provide surgical care during COVID-19, we chose to exclude guidelines and recommendations from this review for two reasons: 1) a high quality review of surgical recommendations for the response to COVID-19 was published by one of the authors just prior to this study^10^ and 2) because there is abundant evidence suggesting guidelines and recommendations for practice are frequently not implemented into clinical practice.^35-41^ Some of the guideline recommendations in the review by Søreide et al.^10^ were implemented within the included studies in the present review; such as creating areas within-hospital for ‘clean’ and ‘contaminated’ cases and workforce redeployment to critical care. However, other recommendations were infrequently noted, such as the dedicated use of isolated, negative pressure ORs for patients with COVID-19. These resource intensive practices may not have been attainable under the pressures of managing public health emergencies and may not be feasible in low-resource settings.

Changes to surgical services, such as cancelling or postponing non-urgent surgeries may be necessary to manage public health emergencies to reduce the risk of contamination and increase capacity within hospitals. However, the impact of these changes remains poorly understood. Many studies reported decreases in surgical volumes, but few other variables were explored with regards to the impact on patients, providers, and healthcare systems. Five studies examined the impact of changes to surgical services among physicians and trainees, and found that training was compromised in some specialties.^18-21^ The finding that medical training was compromised is particularly important for understanding the downstream and long-term repercussions of the response to public health emergencies; decreases in surgical volumes and clinical hours for trainees could have negative and unintended effects on the future quality and safety of patient care.^42^ Studies examining the effects of surgical service alterations on patients noted negative effects on mental health outcomes,^27 28^ pain,^27^ and an increased incidence of death among surgical patients.^22 23 43^

Very few studies described specific actions undertaken to rebuild and resume pre-public health emergencies surgical capacity. This may be due to the fact that most included studies examined the ongoing COVID-19 pandemic, or because few places have implemented specific plans to date. Included studies did describe consideration of system-level factors like availability of PPE and ORs. However, more patient-centric considerations such as organizing child care and requesting time away from their job during a pandemic, are needed. Interestingly, one study reported 14% of surgical patients initiated the cancellation of their surgery,^27^ which suggests patient readiness for surgery during- and post-COVID-19 should be considered. For evidence to inform policy, additional research is needed to understand the impacts of different approaches for resuming surgical services.

This study is, to our knowledge, the first comprehensive scoping review of evidence around reallocation of surgical services during public health emergencies. While this study has several strengths, including a comprehensive search of academic and grey literature sources, and a mix of inductive and deductive data abstraction approaches, there are some limitations that should be considered when interpreting our findings. We modified the Joanna Briggs methodology for scoping reviews,^5^ according to the World Health Organization and Cochrane’s guidance on conducting rapid reviews,^7 8^ with the intent of balancing rigor with a timely and policy-responsive review of the literature. Also, given that the evidence around the COVID-19 pandemic is growing at an unprecedented rate, it is possible that additional studies have been published since we ran our search strategy, especially around resuming surgical services. In order to mitigate this limitation, an ongoing effort to pivot this study into a living review is underway to ensure the data presented is up-to-date. Notably, this review did not identify evidence from any low or middle-income countries who may face unique challenges during a pandemic compared to high income countries described in our review. It is also likely that during the global pandemic, many healthcare institutions have been focused on coping with COVID-19 instead of publishing their experiences; we hope more organizations will add their experience to the literature.

In conclusion, we report early evidence of the operational changes that have occurred internationally in response to public health emergencies which could inform the ongoing response to COVID-19 and future public health emergencies. This study identified a gap in our understanding of the impact of these changes on patients, providers, and the healthcare system which should be the focus of research moving forward to provide an evidence-based approach to managing surgical patients in future public health emergencies.

## Supporting information

PRISMA-ScR checklist

## Data Availability

All data will be available upon reasonable request to the corresponding author.

## Author Contributions

CO contributed to the design and conceptualization of the review, analysis and interpretations of the data, and drafting and revising the manuscript; JSN contributed to the design of the review, interpretation of the data, providing feedback on the manuscript, and gave approval of the final version of the manuscript; AKR contributed to the design of the review, interpretation of the data, providing feedback on the manuscript, and gave approval of the final version of the manuscript; JCD contributed to the interpretation of the data, providing feedback on the manuscript, and gave approval of the final version of the manuscript; JW contributed to the interpretation of the data, providing feedback on the manuscript, and gave approval of the final version of the manuscript; JR contributed to the interpretation of the data, providing feedback on the manuscript, and gave approval of the final version of the manuscript; MB contributed to the interpretation of the data, providing feedback on the manuscript, and gave approval of the final version of the manuscript; KMS contributed to the design and conceptualization of the review, analysis and interpretations of the data, providing feedback on the manuscript, and gave approval of the final version of the manuscript.

