## Appendix A for "Surgery & COVID-19: A rapid scoping review of the impact of COVID-19 on surgical services during public health emergencies"

**Appendix A.** Search Strategy

**Database: Ovid MEDLINE (R) and Epub Ahead of Print, In-Process & Other Non-Indexed Citations and Daily <1946 to May 08, 2020>**
Search Strategy:

1 exp Disease Outbreaks/ (96117)
2 (pandemic* or epidemic* or outbreak* or out break*).mp. (230774)
3 1 or 2 (231940)
4 exp Coronavirus (13456)
5 coronavirus infections/ or severe acute respiratory syndrome/ (11068)
6 Coronaviridae Infections/ (900)
7 coronaviridae/ or coronavirus/ (3916)
8 Influenza a virus, h1n1 subtype/ or Influenza a virus, h3n2 subtype/ or influenza a virus, h5n1 subtype/ (22141)
9 Hemorrahagic Fever, Ebola/ (5316)
10 SARS Virus/ (3038)
11 Middle East Respiratory Syndrome Coronavirus/ (1034)
12 (pneumonia.mp. or exp pneumonia/) and Wuhan.mp. (661)
13 (coronavir* or COVID-19 or SARS-related coronavirus or SARS-CoV-2 or 2019 novel coronavirus or 2019-nCoV or nCoV or h1n1 or h3n2 or h5n1 or avian influenza or avian flu or swine influenza or swine flu or SARS or ebola* or middle east respiratory syndrome or MERS).mp. (77950)
14 (wuhan) adj2 (coronavir* or flu or pneumonia* or COVID-19 or 2019-nCoV).mp. (190)
15 (coronavir*) adj2 (infection*).mp. (8441)
16 4 or 5 or 6 or 7 or 8 or 9 or 10 or 11 or 12 or 13 or 14 or 15 (79915)
17 General Surgery/ (38626)
18 Orthopedic Procedures/ (25371)
19 Traumatology/ (3474)
20 Neurosurgery/ (14892)
21 Obstetrics/ (22533)
22 Anesthesia/ (62587)
23 surgical procedures, operative/ or exp elective surgical procedures/ (68085)
24 exp Arthroplasty, Replacement/ (54362)
25 (general surgery or orthopaedic or orthopedic or trauma or neurosurgery or obstetrics or anesthesia or anaesthesia).mp. (748651)
26 (surger* or operation* or procedure*).mp. (3638036)
27 ((elective or non-urgent) adj2 (surg* or procedure*)).mp. (31794)
28 (surg* adj2 (procedure* or planning or triage or operation* or resource* or backlog or re-organiz* or postpone* or cancel* or capacit* or wait time*)).mp. (444474)
29 ((clinic* or hospital) adj2 (process* or procedure* or triage or planning or performance* or capacit*)).mp. (69185)
30 17 or 18 or 19 or 20 or 21 or 22 or 23 or 24 or 25 or 26 or 27 or 28 or 29 (4101483)
31 3 and 16 and 30 (2075)

**Database: EMBASE <1974 to 2020 May 08>**
Search Strategy:

1 exp epidemic/ (103292)
2 exp pandemic/ (12990)
3 1 or 2 (114458)
4 pandemic influenza/ (4748)
5 exp severe acute respiratory syndrome/ (8505)
6 exp coronaviridae/ or coronaviridae infection/ (14928)
7 coronavirinae/ (2093)
8 Coronavirus infection/ (3397)
9 avian influenza virus/ or "influenza a virus (h1n1)"/ or "influenza a virus (h3n2)"/, "influenza a virus (h5n1)"/ (7466)
10 Ebola hemorrhagic fever/ (5712)
11 exp SARS coronavirus/ (5823)
12 Middle East respiratory syndrome/ (935)
13 (pneumonia.mp. or exp pneumonia/) and Wuhan.mp. (512)
14 (coronavir* or COVID-19 or SARS-related coronavirus or SARS-CoV-2 or 2019 novel coronavirus or 2019-nCoV or nCoV).mp. (92353)
15 (wuhan adj2 (coronavir* or flu or pneumonia* or COVID-19 or 2019-nCoV)).mp. (161)
16 (coronavir* adj2 infection*).mp. (4793)
17 4 or 5 or 6 or 7 or 8 or 9 or 10 or 11 or 12 or 13 or 14 or 15 or 16 (96712)
18 general surgery/ (15070)
19 orthopedic surgery/ (32672)
20 traumatology/ (10653)
21 neurosurgery/ (59847)
22 obstetrics/ (34886)
23 anesthesiological procedure/ (1785)
24 elective surgery/ (34038)
25 replacement arthroplasty/ or exp arthroplasty/ (73583)
26 (general surgery or orthopaedic or orthopedic or trauma or neurosurgery or obstetrics or anaesthesia or anesthesia).mp. (966732)
27 (surger* or operation* or procedure*).mp. (5402047)
28 ((elective or non-urgent adj2 (surg* or procedure*)).mp. (52808)
29 (surg* adj2 (procedure* or planning or triag* or operation* or resource* or backlog or re-organiz* or postpon* or cancel* or capacit* or wait time*)).mp. (187480)
30 ((clinic* or hospital) adj2 (process* or procedure* or triag* or planning or performance* or capacit*)).mp. (83715)
31 18 or 19 or 20 or 21 or 22 or 23 or 24 or 25 or 26 or 27 or 28 or 29 or 30 (5948860)
32 3 and 17 and 31 (1844)

| **Database** | **Pandemic terms**<br>Pandemic, epidemic | **Disease terms**<br>COVID-19, SARS, MERS, pandemic flu (h1n1, h3n2, h5n1), ebola | **Surgical terms**<br>Non-urgent/elective surgery (general, orthopedic, anesthesia, trauma, neurosurgery, obstetrics) |
|---|---|---|---|
| **MEDLINE** | exp Disease Outbreaks/<br>OR<br>(pandemic* or epidemic* or outbreak* or out break*).mp. | exp Coronavirus<br>OR<br>coronavirus infections/ or severe acute respiratory syndrome/<br>OR<br>Coronaviridae Infections/<br>OR<br>coronaviridae/ or coronavirus/<br>OR<br>Influenza a virus, h1n1 subtype/ or Influenza a virus, h3n2 subtype/ or influenza a virus, h5n1 subtype/<br>OR<br>Hemorrahagic Fever, Ebola/<br>OR<br>SARS Virus/<br>OR<br>Middle East Respiratory Syndrome Coronavirus/<br>OR<br>(pneumonia.mp. or exp pneumonia/) and Wuhan.mp.<br>OR<br>(coronavir* or COVID-19 or SARS-related coronavirus or SARS-CoV-2 or 2019 novel coronavirus or 2019-nCoV or nCoV or h1n1 or h3n2 or h5n1 or avian influenza or avian flu or swine influenza or swine flu or SARS or ebola* or middle | General Surgery/<br>OR<br>Orthopedic Procedures/<br>OR<br>Traumatology/<br>OR<br>Neurosurgery/<br>OR<br>Obstetrics/<br>OR<br>Anesthesia/<br>OR<br>surgical procedures, operative/ or exp elective surgical procedures/<br>OR<br>exp Arthroplasty, Replacement/<br>OR<br>(general surgery or orthopaedic or orthopedic or trauma or neurosurgery or obstetrics or anesthesia or anaesthesia).mp.<br>OR<br>(surger* or operation* or procedure*).mp.<br>OR<br>((elective or non-urgent) adj2 (surg* or procedure*)).mp.<br>OR<br>(surg* adj2 (procedure* or planning or triage or operation* or resource* or backlog or re-organiz* or postpone* or cancel* or capacit* or wait time*)).mp. |

| | | | |
|---|---|---|---|
| | | east respiratory syndrome or MERS).mp.<br>OR<br>(wuhan) adj2 (coronavir* or flu or pneumonia* or COVID-19 or 2019-nCoV).mp.<br>OR<br>(coronavir*) adj2 (infection*).mp. | OR<br>((clinic* or hospital) adj2 (process* or procedure* or triage or planning or performance* or capacit*)).mp. |
| **EMBASE** | exp epidemic/<br>OR<br>exp pandemic/ | pandemic influenza/<br>OR<br>exp severe acute respiratory syndrome/<br>OR<br>exp coronaviridae/ or coronaviridae infection/<br>OR<br>coronavirinae/<br>OR<br>Coronavirus infection/<br>OR<br>avian influenza virus/ or "influenza a virus (h1n1)"/ or "influenza a virus (h3n2)"/, "influenza a virus (h5n1)"/<br>OR<br>Ebola hemorrhagic fever/<br>OR<br>exp SARS coronavirus/<br>OR<br>Middle East respiratory syndrome/<br>OR<br>(pneumonia.mp. or exp pneumonia/) and Wuhan.mp.<br>OR<br>(coronavir* or COVID-19 or SARS-related coronavirus or SARS-CoV-2 or 2019 novel coronavirus or 2019-nCoV or nCoV).mp.<br>OR | general surgery/<br>OR<br>orthopedic surgery/<br>OR<br>traumatology/<br>OR<br>neurosurgery/<br>OR<br>obstetrics/<br>OR<br>anesthesiological procedure/<br>OR<br>elective surgery/<br>OR<br>replacement arthroplasty/ or exp arthroplasty/<br>OR<br>(general surgery or orthopaedic or orthopedic or trauma or neurosurgery or obstetrics or anaesthesia or anesthesia).mp.<br>OR<br>(surger* or operation* or procedure*).mp.<br>OR<br>((elective or non-urgent adj2 (surg* or procedure*)).mp.<br>OR<br>(surg* adj2 (procedure* or planning or triag* or operation* or resource* or backlog or re-organiz* or postpon* or cancel* or |

| | | | |
|---|---|---|---|
| | | (wuhan adj2 (coronavir* or flu or pneumonia* or COVID-19 or 2019-nCoV)).mp.<br>OR<br>(coronavir* adj2 infection*).mp. | capacit* or wait time*)).mp.<br>OR<br>((clinic* or hospital) adj2 (process* or procedure* or triag* or planning or performance* or capacit*)).mp. |

**GREY LITERATURE SEARCH PLAN**
Developed in reference to Guide from the University of Toronto (Stapleton, Fuller & Lenton)

STEP 1) Targeted Website Browsing. Will be using any combination of standard terms "coronavirus" OR "Covid-19" OR "Surgery" OR "non-urgent surgery" OR "elective surgery" OR "guidelines")

**NON-GOVERNMENT GROUPS**
World Health Organization (https://www.who.int)
Center for Disease Control and Prevention (https://www.cdc.gov)
European Centre for Disease Prevention and Control (https://www.ecdc.europa.eu/en)

**GOVERNMENTS/HEALTH SYSTEMS**
Canada (https://www.canada.ca/en.html)

- BC (https://www2.gov.bc.ca/gov/content/home)
- AB (https://www.alberta.ca/index.aspx)
- SK (https://www.saskatchewan.ca)
- MB (https://www.gov.mb.ca)
- ON (https://www.ontario.ca/page/government)
- QC (https://www.quebec.ca/en/)
- NB (https://www2.gnb.ca)
- NS (https://beta.novascotia.ca)
- NL & LB (https://www.gov.nl.ca)
- PEI (https://www.princeedwardisland.ca/en)
- Yukon (https://yukon.ca)
- NWT (https://www.gov.nt.ca)
- Nunavut (https://www.gov.nu.ca)

Australia (https://www.australia.gov.au)
Italy (http://www.governo.it)
Singapore (https://www.gov.sg)
China (https://www.gov.cn/english/)
USA (https://www.usa.gov)

**GENERAL SURGICAL GROUPS/COLLEGES**

Royal College of Physicians and Surgeons of Canada (http://www.royalcollege.ca)
American College of Surgeons (https://www.facs.org)
European Surgical Association (https://www.europeansurgicalassociation.org)
College of Surgeons, Singapore (https://www.ams.edu.sg/colleges/CSS/home)
French Surgical Association
German Society of Surgery (https://www.dgch.de/index.php?id=118)
Italian Society of Surgery (SIC) and Italian Association of Hospital Surgeons (ACOI)
Philippine College of Surgeons (https://www.pcs.org.ph)
Royal Australasian College of Surgeons (https://www.surgeons.org)
Royal College of Surgeons of England (https://www.rcseng.ac.uk)
Royal College of Surgeons of Ireland (https://www.rcsi.com/dublin/)
Spanish Society of Surgery (Associacion Espanola de Cirujanos) (https://www.aecirujanos.es)
Swedish Surgical Society (http://www.svenskkirurgi.se)
The Association of Surgeons of South Africa (http://www.surgeon.co.za)
The Pan African Association of Surgeons (http://www.africansurgeons.com)

STEP 2) Advanced Google Searching. Targeting above sites will assess 5 pages of Google Search past last click. Will be using standard and consistent terms “coronavirus” OR “Covid-19” OR “Surgery” OR “non-urgent” OR “guidelines”)

STEP 3) General Search Engine (Google) Search with same terms as above, assessing 5 pages past last click for relevance.

STEP 4) Contact with Knowledge Experts.

| **Source and Site** | **Search Terms** | **Potentially Relevant Results** |
|---|---|---|
| **Health Groups** | | |
| World Health Organization (https://www.who.int, Advanced Search) | [contains all words] elective surgery covid [contains all words] non-urgent surgery covid | 10 |
| Center for Disease Control and Prevention (https://www.cdc.gov) | [contains all words] elective surgery covid [contains all words] non-urgent surgery covid | 8 |
| European Centre for Disease Prevention and Control (https://www.ecdc.europa.eu/en) | [contains all words] elective surgery covid [contains all words] non-urgent surgery covid | 3 |
| **Governments** | | |
| Government of Canada (https://www.canada.ca/en.html) | [contains all words] elective surgery covid | 4 |

| | | |
|---|---|---|
| | [contains all words] non-urgent surgery covid | |
| Government of BC (https://www2.gov.bc.ca/gov) | [contains all words] elective surgery covid [contains all words] non-urgent surgery covid | 2 |
| Government of AB (https://www.alberta.ca) | [contains all words] elective surgery covid [contains all words] non-urgent surgery covid | 2 |
| Government of SK (https://www.saskatchewan.ca) | [contains all words] elective surgery covid [contains all words] non-urgent surgery covid | 6 |
| Government of MB (https://www.gov.mb.ca) | [contains all words] elective surgery covid [contains all words] non-urgent surgery covid | 4 |
| Government of ON (https://www.ontario.ca) | [contains all words] elective surgery covid [contains all words] non-urgent surgery covid | 1 |
| Government of QC (https://www.quebec.ca/en/) | [contains all words] elective surgery covid [contains all words] non-urgent surgery covid | 0 |
| Government of NB (https://www2.gnb.ca) | [contains all words] elective surgery covid [contains all words] non-urgent surgery covid | 1 |
| Government of NS (https://beta.novascotia.ca) | [contains all words] elective surgery covid [contains all words] non-urgent surgery covid | 1 |
| Government of NL & LB (https://www.gov.nl.ca) | [contains all words] elective surgery covid [contains all words] non-urgent surgery covid | 1 |
| Government of PEI (https://www.princeedwardisland.ca) | [contains all words] elective surgery covid | 2 |

| | | |
|---|---|---|
| | [contains all words] non-urgent surgery covid | |
| Government of Yukon (https://yukon.ca) | [contains all words] elective surgery covid [contains all words] non-urgent surgery covid | 1 |
| Government of NWT (https://www.gov.nt.ca) | [contains all words] elective surgery covid [contains all words] non-urgent surgery covid | 0 |
| Government of Nunavut (https://www.gov.nu.ca) | [contains all words] elective surgery covid [contains all words] non-urgent surgery covid | 0 |
| Government of Australia (https://www.australia.gov.au) | [contains all words] elective surgery covid [contains all words] non-urgent surgery covid | 0 |
| Government of Italy (http://www.governo.it) | [contains all words] elective surgery covid [contains all words] non-urgent surgery covid | 0 |
| Government of Singapore (https://www.gov.sg) | [contains all words] elective surgery covid [contains all words] non-urgent surgery covid | 0 |
| Government of China ((https://www.gov.cn/english/) | [contains all words] elective surgery covid [contains all words] non-urgent surgery covid | 0 |
| United States Government (https://www.usa.gov) | [contains all words] elective surgery covid [contains all words] non-urgent surgery covid | 0 |
| Surgical Colleges/Associations | | |
| Royal College of Physicians and Surgeons of Canada (http://www.royalcollege.ca) | [contains all words] elective surgery covid [contains all words] non-urgent surgery covid | 0 |

| | | |
|---|---|---|
| American College of Surgeons (https://www.facs.org) | [contains all words] elective surgery covid [contains all words] non-urgent surgery covid | 23 |
| European Surgical Association (https://www.europeansurgicalassociation.org) | [contains all words] elective surgery covid [contains all words] non-urgent surgery covid | 0 |
| College of Surgeons, Singapore (https://www.ams.edu.sg) | [contains all words] elective surgery covid [contains all words] non-urgent surgery covid | 3 |
| German Society of Surgery (https://www.dgch.de/index.php?id=118) | [contains all words] elective surgery covid [contains all words] non-urgent surgery covid | 0 |
| Philippine College of Surgeons (https://www.pcs.org.ph) | [contains all words] elective surgery covid [contains all words] non-urgent surgery covid | 11 |
| Royal Australasian College of Surgeons (https://www.surgeons.org) | [contains all words] elective surgery covid [contains all words] non-urgent surgery covid | 12 |
| Royal College of Surgeons of England (https://www.rcseng.ac.uk) | [contains all words] elective surgery covid [contains all words] non-urgent surgery covid | 7 |
| Royal College of Surgeons of Ireland (https://www.rcsi.com/dublin/) | [contains all words] elective surgery covid [contains all words] non-urgent surgery covid | 8 |
| Spanish Society of Surgery (Associacion Espanola de Cirujanos) (https://www.aecirujanos.es) | [contains all words] elective surgery covid [contains all words] non-urgent surgery covid | 1 |
| Swedish Surgical Society (http://www.svenskkirurgi.se) | [contains all words] elective surgery covid [contains all words] non-urgent surgery covid | 0 |

| | | |
|---|---|---|
| The Association of Surgeons of South Africa (http://www.surgeon.co.za) | [contains all words] elective surgery covid [contains all words] non-urgent surgery covid | 0 |
| The Pan African Association of Surgeons (http://www.africansurgeons.com) | [contains all words] elective surgery covid [contains all words] non-urgent surgery covid | 0 |
| | Total | 111 |
