## Appendix B for "Surgery & COVID-19: A rapid scoping review of the impact of COVID-19 on surgical services during public health emergencies"

**Appendix B.** Data Abstraction Form

This form has been developed by adopting and customizing the 'Data collection form- RCTs and NRS' produced by The Cochrane Collaboration. Customization includes the addition of new sections, as well as the omission of sections not relevant to the review.

### Notes on using data extraction form:

- Be consistent in the order and style you use to describe the information for each report.
- Record any missing information as unclear or not described, to make it clear that the information was not found in the study report(s), not that you forgot to extract it.
- Include any instructions and decision rules on the data collection form, or in an accompanying document. It is important to practice using the form and give training to any other authors using the form.

#### 1. General Information

| | |
|---|---|
| 1. Date form completed (dd/mm/yyyy) | / / |
| 2. Name of person extracting data | ☐ Connor M. ORielly<br>☐ Khara M. Sauro |
| 3. Contact details of person extracting data | |
| 4. Title of Article/Abstract (that data are extracted from) | |
| 5. Study ID (plus surname of author and year of study publication) | |
| 6. Study country of origin | |
| 7. Study funding source | |
| 8. Possible conflicts of interest | ☐ Reported<br>☐ Not Reported |
| 9. Notes: | |

#### 2. Eligibility

| Study Characteristics | Inclusion Criteria | Location in text (page#/fig#/table#) |
|---|---|---|
| 10. Type of study (design) | ☐ Case Study ☐Case Series<br>☐ Observational | |

| | |
|---|---|
| 11. Population description | |
| 12. Focused diseases/conditions | |
| 13. Types of outcome measures | |
| 14. Decision (with reasons for either inclusion or exclusion) | ☐ Include<br>☐ Exclude<br>If Exclude, explain: |
| 15. Notes | |

**IF STUDY IS EXCLUDED FROM REVIEW, DO NOT CONTINUE**

**2. Population and setting**

| | Description | Location in text (page#/fig#/table#) |
|---|---|---|
| 16. Population description | | |
| 17. Source/setting of the population (e.g., urban, rural, ethnic group) | | |
| 18. Method(s) of recruitment of participants | ☐ Random<br>☐ Non-Random<br><br>Method: | |
| 19. Notes: | | |

**3. Methods**

| | Descriptions as stated in the article/abstract | Location in text (page#/fig#/table#) |
|---|---|---|

| | |
|---|---|
| 20. Aim of study | |
| 21. Design (e.g., cross-sectional, RCT, CCT) | ☐ Case Study ☐ Case Series<br>☐ Observational |
| 22. Sampling technique (e.g., random or convenience) | ☐ Random<br>☐ Non-Random |
| 23. Study start date (dd/mm/yyyy) | / / |
| 24. Study end date/duration (dd/mm/yyyy) | / /<br>**OR** Duration: |
| 25. Notes: | |

**4. Participants**

| | Descriptions as stated in the article/abstract | Location in text (page#/fig#/table#) |
|---|---|---|
| 26. Total number of participants/Sample size | | |
| 27. Age | | |
| 28. Sex | ☐ Both Sexes<br>☐ Males<br>☐ Females | |
| 29. Genders Represented | | |
| 30. Country | | |
| 31. Predominant medical complaint | | |
| 32. Co-morbidities (if any) | | |
| 33. Definition of 'Frequent use' (if any) | | |

| 34. Notes: |
| --- |

**5. Outcomes**

*Keep only the tables for outcomes relevant to this particular study, duplicate tables as necessary.*

| Outcome 1: Alterations to surgical programming (RQ1) | Descriptions as stated in the article/abstract | Location in text (page#/fig#/table#) |
| --- | --- | --- |
| 35. Outcome definition | | |
| 36. Time points measured | | |
| 37. Domains of change | ☐ Surgical triage<br>☐ PPE<br>☐ Workforce<br>☐ Patient care<br>☐ Environmental | |
| 38. Unit of measurement | ☐ Group<br>☐ Individual | |
| 39. Notes: | | |

| Outcome 2: Patient-level outcomes (RQ2) | Descriptions as stated in the article/abstract | Location in text (page#/fig#/table#) |
| --- | --- | --- |
| 40. Specify Outcome | | |
| 41. Outcome definition | | |
| 42. Time points measured | | |
| 43. Unit of Measurement | ☐ Group<br>☐ Individual | |
| 44. Unit of measurement | | |
| 45. Notes: | | |

| Outcome 3: System-level outcomes (RQ2) | Descriptions as stated in the article/abstract | Location in text (page#/fig#/table#) |
| --- | --- | --- |
| 46. Specify Outcome | | |
| 47. Outcome definition | | |
| 48. Time points measured | | |
| 49. Unit of Measurement | ☐ Group<br>☐ Individual | |
| 50. Unit of measurement | | |
| 51. Notes: | | |

| Outcome 4: Practitioner-level outcomes (RQ2) | Descriptions as stated in the article/abstract | Location in text (page#/fig#/table#) |
| --- | --- | --- |
| 52. Specify Outcome | | |
| 53. Outcome definition | | |
| 54. Time points measured | | |
| 55. Unit of Measurement | ☐ Group<br>☐ Individual | |
| 56. Unit of measurement | | |
| 57. Notes: | | |

| Outcome 5: Alterations to rebuild capacity (RQ3) | Descriptions as stated in the article/abstract | Location in text (page#/fig#/table#) |
| --- | --- | --- |
| 58. Outcome definition | | |
| 59. Time points measured | | |

| | |
|---|---|
| 60. Domains of change | ☐ Surgical triage<br>☐ PPE<br>☐ Workforce<br>☐ Patient care<br>☐ Environmental |
| 61. Unit of measurement | ☐ Group<br>☐ Individual |
| 62. Notes: | |

**6. Results and findings**

*Keep only the tables for outcomes relevant to this particular study, duplicate tables as necessary.*

| Outcome 1: Alterations to surgical programming (RQ1) | Descriptions as stated in the article/abstract | Location in text (page#/fig#/table#) |
|---|---|---|
| 63. Subgroup (if applicable, e.g., age/sex specific reporting) | | |
| 64. Results | | |
| 65. Response/non-response rate | | |
| 66. Unit of analysis (i.e., individual or group) | | |
| 67. Notes: | | |

| Outcome 2: Patient-level outcomes (RQ2) | Descriptions as stated in the article/abstract | Location in text (page#/fig#/table#) |
|---|---|---|
| 68. Subgroup (if applicable, e.g., age/sex specific reporting) | | |
| 69. Results | | |
| 70. Response/non-response rate | | |
| 71. Unit of analysis (i.e., individual or group) | | |
| 72. Notes: | | |

| Outcome 3: System-level outcomes (RQ2) | Descriptions as stated in the article/abstract | Location in text (page#/fig#/table#) |
|---|---|---|
| 73. Subgroup (if applicable, e.g., age/sex specific reporting) | | |
| 74. Results | | |
| 75. Response/non-response rate | | |
| 76. Unit of analysis (i.e., individual or group) | | |
| 77. Notes: | | |

| Outcome 4: Practitioner-level outcomes (RQ2) | Descriptions as stated in the article/abstract | Location in text (page#/fig#/table#) |
|---|---|---|
| 78. Subgroup (if applicable, e.g., age/sex specific reporting) | | |
| 79. Results | | |
| 80. Response/non-response rate | | |
| 81. Unit of analysis (i.e., individual or group) | | |
| 82. Notes: | | |

| Outcome 5: Alterations to rebuild capacity (RQ3) | Descriptions as stated in the article/abstract | Location in text (page#/fig#/table#) |
|---|---|---|
| 83. Subgroup (if applicable, e.g., age/sex specific reporting) | | |
| 84. Results | | |
| 85. Response/non-response rate | | |
| 86. Unit of analysis (i.e., individual or group) | | |
| 87. Notes: | | |

**Research Questions informed by this study *(tick only boxes matching the outcome tables kept above)***

☐ Changes to surgical programming in response to public health emergency

☐ Impacts of changes to surgical programming
☐ Actions to rebuild surgical capacity post-public health emergency

**7. Strengths, limitations and mitigation strategy**

| | Descriptions as stated in the article/abstract | Location in text (page#/fig#/table#) |
|---|---|---|
| 88. Strengths | | |
| 89. Limitations | | |
| 90. Strategies to overcome limitations | | |
| 91. Notes: | | |

**8. Conclusion and other information**

| | Descriptions as stated in the article/abstract | Location in text (page#/fig#/table#) |
|---|---|---|
| 92. Key conclusions of study authors | | |
| 93. Notes: | | |
