## Appendix C for "Surgery & COVID-19: A rapid scoping review of the impact of COVID-19 on surgical services during public health emergencies"

**Appendix C.** Complete list of references for included studies

44. Gupta A, Arora V, Nair D, et al. Status and strategies for the management of head and neck cancer during COVID-19 pandemic: Indian scenario. *Head Neck* 2020 doi: 10.1002/hed.26227 [published Online First: 2020/05/02]
45. Haines CJ, Chu YW, Chung TK. The effect of Severe Acute Respiratory Syndrome on a hospital obstetrics and gynaecology service. *BJOG* 2003;110(7):643-5. doi: http://dx.doi.org/10.1016/S1470-0328%2803%2903007-6 [published Online First: 2003/07/05]
46. British Columbia Ministry of Health. A Commitment to Surgical Renewal in BC. Canada, 2020.
47. Hemingway JF, Singh N, Starnes BW. Emerging practice patterns in vascular surgery during the COVID-19 pandemic. *J Vasc Surg* 2020 doi: 10.1016/j.jvs.2020.04.492 [published Online First: 2020/05/04]
48. Hormati A, Ghadir MR, Zamani F, et al. Preventive strategies used by GI physicians during the COVID-19 pandemic. *New Microbes New Infect* 2020;35:100676. doi: 10.1016/j.nmni.2020.100676 [published Online First: 2020/04/16]
49. Hu Y-J, Zhang J-M, Chen Z-P. Experiences of practicing surgical neuro-oncology during the COVID-19 pandemic. *Journal of neuro-oncology* 2020;148(1):199-200. doi: 10.1007/s11060-020-03489-6 [published Online First: 2020/04/10]
50. Jean WC, Ironside NT, Sack KD, et al. The impact of COVID-19 on neurosurgeons and the strategy for triaging non-emergent operations: a global neurosurgery study. *Acta Neurochir (Wien)* 2020;162(6):1229-40. doi: 10.1007/s00701-020-04342-5 [published Online First: 2020/04/22]
51. Kempa M, Gulaj M, Farkowski MM, et al. Electrotherapy and electrophysiology procedures during the coronavirus disease 2019 pandemic: an opinion of the Heart Rhythm Section of the Polish Cardiac Society (with an update). *Kardiol Pol* 2020;78(5):488-92. doi: 10.33963/KP.15338 [published Online First: 2020/05/06]
52. Kessler RA, Zimering J, Gilligan J, et al. Neurosurgical management of brain and spine tumors in the COVID-19 era: an institutional experience from the epicenter of the pandemic. *J Neurooncol* 2020 doi: 10.1007/s11060-020-03523-7 [published Online First: 2020/05/07]
53. Konda SR, Dankert JF, Merkow D, et al. COVID-19 Response in the Global Epicenter: Converting a New York City Level 1 Orthopedic Trauma Service into a Hybrid Orthopedic and Medicine COVID-19 Management Team. *J Orthop Trauma* 2020 doi: 10.1097/BOT.0000000000001792 [published Online First: 2020/05/02]
54. Kuo IC, Pellegrino F, Fornero P, et al. H1N1 pandemic and ophthalmology. *Ophthalmology* 2010;117(2):405. doi: 10.1016/j.ophtha.2009.11.035 [published Online First: 2010/02/10]
55. Lai THT, Tang EWH, Chau SKY, et al. Stepping up infection control measures in ophthalmology during the novel coronavirus outbreak: an experience from Hong Kong. *Graefes Arch Clin Exp Ophthalmol* 2020;258(5):1049-55. doi: 10.1007/s00417-020-04641-8 [published Online First: 2020/03/04]
56. Lancaster EM, Sosa JA, Sammann A, et al. Rapid Response of an Academic Surgical Department to the COVID-19 Pandemic: Implications for Patients, Surgeons, and the Community. *J Am Coll Surg* 2020;230(6):1064-73. doi: 10.1016/j.jamcollsurg.2020.04.007 [published Online First: 2020/04/13]
57. Langer PD, Bernardini FP. Oculofacial Plastic Surgery and the COVID-19 Pandemic: Current Reactions and Implications for the Future. *Ophthalmology* 2020 doi: 10.1016/j.ophtha.2020.04.035 [published Online First: 2020/04/30]

58. Lauterio A, De Carlis R, Belli L, et al. How to guarantee liver transplantation in the north of Italy during the COVID-19 pandemic: A sound transplant protection strategy. *Transpl Int* 2020 doi: 10.1111/tri.13633 [published Online First: 2020/04/30]
59. Lee AKF, Cho RHW, Lau EHL, et al. Mitigation of head and neck cancer service disruption during COVID-19 in Hong Kong through telehealth and multi-institutional collaboration. *Head Neck* 2020 doi: 10.1002/hed.26226 [published Online First: 2020/05/02]
60. Leong Tan GW, Chandrasekar S, Lo ZJ, et al. Early experience in the COVID-19 pandemic from a vascular surgery unit in a Singapore tertiary hospital. *J Vasc Surg* 2020 doi: 10.1016/j.jvs.2020.04.014 [published Online First: 2020/04/20]
61. Li Y, Yang N, Li X, et al. Strategies for prevention and control of the 2019 novel coronavirus disease in the department of kidney transplantation. *Transpl Int* 2020 doi: 10.1111/tri.13634 [published Online First: 2020/05/02]
62. Liebensteiner MC, Khosravi I, Hirschmann MT, et al. Massive cutback in orthopaedic healthcare services due to the COVID-19 pandemic. *Knee Surg Sports Traumatol Arthrosc* 2020;28(6):1705-11. doi: 10.1007/s00167-020-06032-2 [published Online First: 2020/05/02]
63. Liu EH, Koh KF, Chen FG. Outbreak of severe acute respiratory syndrome in Singapore and modifications in the anesthesia service. *Anesthesiology* 2004;100(6):1629-30. doi: 10.1097/00000542-200406000-00062 [published Online First: 2004/05/29]
64. Mak ST, Yuen HK. Oculoplastic surgery practice during the COVID-19 novel coronavirus pandemic: experience sharing from Hong Kong. *Orbit* 2020:1-3. doi: 10.1080/01676830.2020.1754435 [published Online First: 2020/04/17]
65. Marti C, Sanchez-Mendez JI. Neoadjuvant endocrine therapy for luminal breast cancer treatment: a first-choice alternative in times of crisis such as the COVID-19 pandemic. *Ecancermedicalscience* 2020;14:1027. doi: 10.3332/ecancer.2020.1027 [published Online First: 2020/05/06]
66. Maurizi G, Rendina EA. A High-Volume Thoracic Surgery Division into the Storm of the Covid-19 Pandemic. *Ann Thorac Surg* 2020 doi: 10.1016/j.athoracsur.2020.03.015 [published Online First: 2020/04/15]
67. McBride KE, Brown KGM, Fisher OM, et al. Impact of the COVID-19 pandemic on surgical services: early experiences at a nominated COVID-19 centre. *ANZ J Surg* 2020;90(5):663-65. doi: 10.1111/ans.15900 [published Online First: 2020/04/08]
68. McMillan T, Williamson K. Resuming non-urgent surgeries and allied health services, 2020.
69. Meneghini RM. Resource Reallocation during the COVID-19 Pandemic in a Suburban Hospital System: Implications for Outpatient Hip and Knee Arthroplasty. *J Arthroplasty* 2020 doi: 10.1016/j.arth.2020.04.051 [published Online First: 2020/05/08]
70. Meyer M, Prost S, Farah K, et al. Spine Surgical Procedures during Coronavirus Disease 2019 Pandemic: Is It Still Possible to Take Care of Patients? Results of an Observational Study in the First Month of Confinement. *Asian Spine J* 2020 doi: 10.31616/asj.2020.0197 [published Online First: 2020/05/10]
71. Morgan C, Ahluwalia AK, Aframian A, et al. The impact of the novel coronavirus on trauma and orthopaedics in the UK. *Br J Hosp Med (Lond)* 2020;81(4):1-6. doi: 10.12968/hmed.2020.0137 [published Online First: 2020/04/29]
72. Nair AG, Gandhi RA, Natarajan S. Effect of COVID-19 related lockdown on ophthalmic practice and patient care in India: Results of a survey. *Indian J Ophthalmol* 2020;68(5):725-30. doi: 10.4103/ijo.IJO_797_20 [published Online First: 2020/04/23]

73. Nassar AH, Zern NK, McIntyre LK, et al. Emergency Restructuring of a General Surgery Residency Program During the Coronavirus Disease 2019 Pandemic: The University of Washington Experience. *JAMA Surg* 2020 doi: 10.1001/jamasurg.2020.1219 [published Online First: 2020/04/07]
74. Park J, Yoo SY, Ko JH, et al. Infection Prevention Measures for Surgical Procedures during a Middle East Respiratory Syndrome Outbreak in a Tertiary Care Hospital in South Korea. *Sci Rep* 2020;10(1):325. doi: 10.1038/s41598-019-57216-x [published Online First: 2020/01/17]
75. Park JS, El-Sayed IH, Young VN, et al. Development of clinical care guidelines for faculty and residents in the era of COVID-19. *Head Neck* 2020 doi: 10.1002/hed.26225 [published Online First: 2020/04/30]
76. Patel RJ, Kejner A, McMullen C. Early institutional head and neck oncologic and microvascular surgery practice patterns across the United States during the SARS-CoV-2 (COVID19) pandemic. *Head Neck* 2020;42(6):1168-72. doi: 10.1002/hed.26189 [published Online First: 2020/04/25]
77. Patel ZM, Fernandez-Miranda J, Hwang PH, et al. Letter: Precautions for Endoscopic Transnasal Skull Base Surgery During the COVID-19 Pandemic. *Neurosurgery* 2020 doi: 10.1093/neuros/nyaa125 [published Online First: 2020/04/16]
78. Pelt CE, Campbell KL, Gililland JM, et al. The Rapid Response to the COVID-19 Pandemic by the Arthroplasty Divisions at Two Academic Referral Centers. *J Arthroplasty* 2020 doi: 10.1016/j.arth.2020.04.030 [published Online First: 2020/05/02]
79. Pittet D. Hopitaux Universitaire de Genève (HUG)- A model hospital for COVID-19 patient management, 2020.
80. Prachand VN, Milner R, Angelos P, et al. Medically Necessary, Time-Sensitive Procedures: Scoring System to Ethically and Efficiently Manage Resource Scarcity and Provider Risk During the COVID-19 Pandemic. *J Am Coll Surg* 2020 doi: 10.1016/j.jamcollsurg.2020.04.011 [published Online First: 2020/04/13]
81. Price KN, Thiede R, Shi VY, et al. Strategic dermatology clinical operations during the coronavirus disease 2019 (COVID-19) pandemic. *J Am Acad Dermatol* 2020;82(6):e207-e09. doi: 10.1016/j.jaad.2020.03.089 [published Online First: 2020/04/12]
82. Qadan M, Hong TS, Tanabe KK, et al. A Multidisciplinary Team Approach for Triage of Elective Cancer Surgery at the Massachusetts General Hospital During the Novel Coronavirus COVID-19 Outbreak. *Ann Surg* 2020 doi: 10.1097/SLA.0000000000003963 [published Online First: 2020/04/18]
83. Ralli M, Greco A, de Vincentiis M. The Effects of the COVID-19/SARS-CoV-2 Pandemic Outbreak on Otolaryngology Activity in Italy. *Ear Nose Throat J* 2020:145561320923893. doi: 10.1177/0145561320923893 [published Online First: 2020/04/30]
84. Rampinelli V, Mattavelli D, Gualtieri T, et al. Reshaping head and neck reconstruction policy during the COVID-19 pandemic peak: Experience in a front-line institution. *Auris Nasus Larynx* 2020 doi: 10.1016/j.anl.2020.04.008 [published Online First: 2020/05/05]
85. Randelli PS, Compagnoni R. Management of orthopaedic and traumatology patients during the Coronavirus disease (COVID-19) pandemic in northern Italy. *Knee Surg Sports Traumatol Arthrosc* 2020;28(6):1683-89. doi: 10.1007/s00167-020-06023-3 [published Online First: 2020/04/27]

101. Sun Y, Mao Y. Editorial. Response to COVID-19 in Chinese neurosurgery and beyond. *Journal of neurosurgery* 2020:1-2. doi: 10.3171/2020.3.JNS20929
102. National Clinical Programme in Surgery. Surgical Patient Flow During COVID-19 Pandemic, 2020.
103. Tan TK. How severe acute respiratory syndrome (SARS) affected the department of anaesthesia at Singapore General Hospital. *Anaesth Intensive Care* 2004;32(3):394-400. doi: 10.1177/0310057x0403200316 [published Online First: 2004/07/22]
104. Tan YQ, Wu QH, Chiong E. Preserving Operational Capability While Building Capacity During the COVID-19 Pandemic: A Tertiary Urology Centre's Experience. *Urology* 2020 doi: 10.1016/j.urology.2020.04.079 [published Online First: 2020/05/04]
105. Tan YT, Wang JW, Zhao K, et al. Preliminary Recommendations for Surgical Practice of Neurosurgery Department in the Central Epidemic Area of 2019 Coronavirus Infection. *Curr Med Sci* 2020;40(2):281-84. doi: 10.1007/s11596-020-2173-5 [published Online First: 2020/03/29]
106. Tay K, Kamarul T, Lok WY, et al. COVID-19 in Singapore and Malaysia: Rising to the Challenges of Orthopaedic Practice in an Evolving Pandemic. *Malays Orthop J* 2020;14(2) doi: 10.5704/MOJ.2007.001 [published Online First: 2020/04/22]
107. Tay KJD, Lee YHD. Trauma and orthopaedics in the COVID-19 pandemic: breaking every wave. *Singapore Med J* 2020 doi: 10.11622/smedj.2020063 [published Online First: 2020/04/22]
108. Thaler M, Khosravi I, Hirschmann MT, et al. Disruption of joint arthroplasty services in Europe during the COVID-19 pandemic: an online survey within the European Hip Society (EHS) and the European Knee Associates (EKA). *Knee Surg Sports Traumatol Arthrosc* 2020;28(6):1712-19. doi: 10.1007/s00167-020-06033-1 [published Online First: 2020/05/04]
109. Tolone S, Gambardella C, Brusciano L, et al. Telephonic triage before surgical ward admission and telemedicine during COVID-19 outbreak in Italy. Effective and easy procedures to reduce in-hospital positivity. *Int J Surg* 2020;78:123-25. doi: 10.1016/j.ijsu.2020.04.060 [published Online First: 2020/05/04]
110. Too CW, Wen DW, Patel A, et al. Interventional Radiology Procedures for COVID-19 Patients: How we Do it. *Cardiovasc Intervent Radiol* 2020;43(6):827-36. doi: 10.1007/s00270-020-02483-9 [published Online First: 2020/04/29]
111. Topf MC, Shenson JA, Holsinger FC, et al. Framework for prioritizing head and neck surgery during the COVID-19 pandemic. *Head Neck* 2020;42(6):1159-67. doi: 10.1002/hed.26184 [published Online First: 2020/04/17]
112. Tsui KL, Li SK, Li MC, et al. Preparedness of the cardiac catheterization laboratory for severe acute respiratory syndrome (SARS) and other epidemics. *J Invasive Cardiol* 2005;17(3):149-52. [published Online First: 2005/05/04]
113. Tzeng C-WD, Tran Cao HS, Roland CL, et al. Surgical decision-making and prioritization for cancer patients at the onset of the COVID-19 pandemic: A multidisciplinary approach. *Surgical Oncology* 2020;34:182-85. doi: 10.1016/j.suronc.2020.04.029
114. Unal EU, Mavioglu HL, Iscan HZ. Vascular surgery in the COVID-19 pandemic. *J Vasc Surg* 2020 doi: 10.1016/j.jvs.2020.04.480 [published Online First: 2020/04/29]
115. Vaccaro AR, Getz CL, Cohen BE, et al. Practice Management During the COVID-19 Pandemic. *J Am Acad Orthop Surg* 2020;28(11):464-70. doi: 10.5435/JAAOS-D-20-00379 [published Online First: 2020/04/15]

116. Valenza F, Papagni G, Marchiano A, et al. Response of a comprehensive cancer center to the COVID-19 pandemic: the experience of the Fondazione IRCCS-Istituto Nazionale dei Tumori di Milano. *Tumori* 2020:300891620923790. doi: 10.1177/0300891620923790 [published Online First: 2020/05/05]
117. van de Haar J, Hoes LR, Coles CE, et al. Caring for patients with cancer in the COVID-19 era. *Nat Med* 2020;26(5):665-71. doi: 10.1038/s41591-020-0874-8 [published Online First: 2020/05/15]
118. Vicini E, Galimberti V, Naninato P, et al. COVID-19: The European institute of oncology as a "hub" centre for breast cancer surgery during the pandemic in Milan (Lombardy region, northern Italy) - A screenshot of the first month. *Eur J Surg Oncol* 2020 doi: 10.1016/j.ejso.2020.04.026 [published Online First: 2020/05/04]
119. Vlantis AC, Tsang RK, Wong DK, et al. The impact of severe acute respiratory syndrome on otorhinolaryngological services at the Prince of Wales Hospital in Hong Kong. *Laryngoscope* 2004;114(1):171-4. doi: 10.1097/00005537-200401000-00032 [published Online First: 2004/01/08]
120. Walker JP. Resuming Elective Surgery at UTMB Predicated on Patient and Staff Well-Being, 2020.
121. Wan IYP, Wat KHY, Ng CSH, et al. Evaluation of the emotional status of patients on a waiting list for thoracic surgery during the outbreak of Severe Acute Respiratory Syndrome(SARS). *Stress and Health* 2004;20(4):209-12. doi: 10.1002/smi.1013
122. Wang X, Wang Z, Yao C, et al. Management of ophthalmic perioperative period during 2019 novel coronavirus disease outbreak. *Zhonghua Shiyan Yanke Zazhi/Chinese Journal of Experimental Ophthalmology* 2020;38(3):200-03. doi: http://dx.doi.org/10.3760/cma.j.issn115989-20200224-00100
123. Wasser LM, Assayag E, Tsessler M, et al. Response of ophthalmologists in Israel to the novel coronavirus (2019-nCoV) outbreak. *Graefes Arch Clin Exp Ophthalmol* 2020 doi: 10.1007/s00417-020-04694-9 [published Online First: 2020/04/30]
124. Williams AM, Kalra G, Commiskey PW, et al. Ophthalmology Practice During the Coronavirus Disease 2019 Pandemic: The University of Pittsburgh Experience in Promoting Clinic Safety and Embracing Video Visits. *Ophthalmol Ther* 2020:1-9. doi: 10.1007/s40123-020-00255-9 [published Online First: 2020/05/08]
125. Wong J, Goh QY, Tan Z, et al. Preparing for a COVID-19 pandemic: a review of operating room outbreak response measures in a large tertiary hospital in Singapore. *Canadian Journal of Anesthesia/Journal canadien d'anesthésie* 2020;67(6):732-45. doi: 10.1007/s12630-020-01620-9
126. Wu V, Noel CW, Forner D, et al. Considerations for head and neck oncology practices during the coronavirus disease 2019 (COVID-19) pandemic: Wuhan and Toronto experience. *Head Neck* 2020;42(6):1202-08. doi: 10.1002/hed.26205 [published Online First: 2020/04/28]
127. Zangrillo A, Beretta L, Silvani P, et al. Fast reshaping of intensive care unit facilities in a large metropolitan hospital in Milan, Italy: facing the COVID-19 pandemic emergency. *Crit Care Resusc* 2020 [published Online First: 2020/04/02]
128. Zarzaur BL, Stahl CC, Greenberg JA, et al. Blueprint for Restructuring a Department of Surgery in Concert With the Health Care System During a Pandemic: The University of Wisconsin Experience. *JAMA Surg* 2020 doi: 10.1001/jamasurg.2020.1386 [published Online First: 2020/04/15]

129. Zeng L, Su T, Huang L. Strategic plan for management in oral and maxillofacial surgery during COVID-19 epidemic. *Oral Oncol* 2020;105:104715. doi: 10.1016/j.oraloncology.2020.104715 [published Online First: 2020/04/20]
130. Zhen L, Lin T, Zhao ML, et al. [Management strategy for the resumption of regular diagnosis and treatment in gastrointestinal surgery department during the outbreak of coronavirus disease 2019 (COVID-19)]. *Zhonghua Wei Chang Wai Ke Za Zhi* 2020;23(4):321-26. doi: 10.3760/cma.j.issn.1671-0274.2020-0316-00146 [published Online First: 2020/04/21]
131. Zizzo M, Bollino R, Castro Ruiz C, et al. Surgical management of suspected or confirmed SARS-CoV-2 (COVID-19)-positive patients: a model stemming from the experience at Level III Hospital in Emilia-Romagna, Italy. *Eur J Trauma Emerg Surg* 2020 doi: 10.1007/s00068-020-01377-2 [published Online First: 2020/04/30]
132. Zoia C, Bongetta D, Veiceschi P, et al. Neurosurgery during the COVID-19 pandemic: update from Lombardy, northern Italy. *Acta Neurochir (Wien)* 2020;162(6):1221-22. doi: 10.1007/s00701-020-04305-w [published Online First: 2020/03/31]
